# A methodology of phenotyping ICU patients from EHR data: high-fidelity, personalized, and interpretable phenotypes estimation

**DOI:** 10.1101/2023.03.15.23287315

**Authors:** Yanran Wang, J.N. Stroh, George Hripcsak, Cecilia C. Low Wang, Tellen D. Bennett, Julia Wrobel, Caroline Der Nigoghossian, Scott Mueller, Jan Claassen, D.J. Albers

## Abstract

**Objective:** Computing phenotypes that provide high-fidelity, time-dependent characterizations and yield personalized interpretations is challenging, especially given the complexity of physiological and healthcare systems and clinical data quality. This paper develops a methodological pipeline to estimate unmeasured physiological parameters and produce high-fidelity, personalized phenotypes anchored to physiological mechanics from electronic health record (EHR).

**Methods:** A methodological phenotyping pipeline is developed that computes new phenotypes defined with unmeasurable computational biomarkers quantifying specific physiological properties in real time. Working within the inverse problem framework, this pipeline is applied to the glucose-insulin system for ICU patients using data assimilation to estimate an established mathematical physiological model with stochastic optimization. This produces physiological model parameter vectors of clinically unmeasured endocrine properties, here insulin secretion, clearance, and resistance, estimated for individual patient. These physiological parameter vectors are used as inputs to unsupervised machine learning methods to produce phenotypic labels and discrete physiological phenotypes. These phenotypes are inherently interpretable because they are based on parametric physiological descriptors. To establish potential clinical utility, the computed phenotypes are evaluated with external EHR data for consistency and reliability and with clinician face validation.

**Results:** The phenotype computation was performed on a cohort of 109 ICU patients who received no or short-acting insulin therapy, rendering continuous and discrete physiological phenotypes as specific computational biomarkers of unmeasured insulin secretion, clearance, and resistance on time windows of three days. Six, six, and five discrete phenotypes were found in the first, middle, and last three-day periods of ICU stays, respectively. Computed phenotypic labels were predictive with an average accuracy of 89%. External validation of discrete phenotypes showed coherence and consistency in clinically observable differences based on laboratory measurements and ICD 9/10 codes and clinical concordance from face validity. A particularly clinically impactful parameter, insulin secretion, had a concordance accuracy of 83%*±*27%.

**Conclusion:** The new physiological phenotypes computed with individual patient ICU data and defined by estimates of mechanistic model parameters have high physiological fidelity, are continuous, time-specific, personalized, interpretable, and predictive. This methodology is generalizable to other clinical and physiological settings and opens the door for discovering deeper physiological information to personalize medical care.

## 1 Introduction

Characterizing humans with quantifiable properties in a specific context, embodied in the concept of phenotypes, is essential to medical practice and research. However, computing new phenotypes composed of physiological information not directly in electronic health record (EHR) that are potentially therapeutically informative or actionable for, e.g., personalizing treatment, is often difficult. Here we undertake the challenge of inferring unmeasured physiological parameters to produce high-fidelity phenotypes anchored to physiological mechanics. To achieve this goal, we construct a methodological pipeline that will generalize to larger cohorts and other physiological settings, to produce personalized and interpretable information in real time.

### Background phenotyping efforts for knowledge extraction

Computable phenotyping methods employ machine learning (ML) techniques to infer latent or otherwise unobservable properties from observations in EHR with consistent efficiency [1][2][3][4][5][6][7]. These ML-based phenotyping methods can produce reliable and reproducible knowledge extraction but can still suffer from EHR data complexities. Specifically, high-dimensional EHR data [8][9][10][11], with inconsistent quality from disparate data systems [12], are highly complex and have data sparsity and quality issues. These complexities are complicated by inherent recording biases [8][10] in the health care process [13]. These issues lead to accurate and robust phenotyping methods tending to be work-intensive and time-consuming [8][14][15]. Moreover, the knowledge-based temporal abstraction and ML-based methods [16][17] tend to not incorporate external information or knowledge, leading to broadly-defined phenotypes that contain implicit information from temporal representation mining the health care process recorded in EHR.

### Phenotyping as an inverse problem

The phenotyping knowledge extraction problem with clinical data in EHR can also be seen as a classic inverse problem [18][19][20][21]. This paradigm begins by calculating from explicitly measurable data, e.g., blood glucose, the unmeasurable but implicitly quantifiable properties, e.g., insulin secretion, by estimating a model of glucose-insulin mechanics [22][23][24][25]. Usually, but not always, this nonlinear model consists of a set of ordinary differential equations that represent our mathematized knowledge of a physical system, e.g., a mathematical physiological model [26]. This model has states that evolve fast and continuously in time while being constrained by model structure and parameters that define the slow-evolving underlying variables, e.g., insulin secretion, which can change only when the model is estimated. Ideally, we estimate model states and parameters with measurements that correspond to a complete set of states and parameters. In real clinical settings, though, only small portions of model states are observable and measured, such as blood glucose, insulin administration, etc. In such case, the inverse problem framework reverses a typical forward problem [19] or is the process of estimating variables of underlying mechanics believed to be causal from data. Under this framework, we estimate from measurements the unobservable model parameters, states, and sometimes initial conditions. Essentially, this model estimation is an optimization process for computationally tuning unobservable model parameters and states to minimize the error between observations, such as blood glucose measurements, and their counterparts of continuously simulated model states. This estimation of unobservable properties is one of our goals in using the inverse problem framework for phenotyping knowledge extraction.

### Inverse problems compared with ML

What differentiates the inverse problem framework from ML are the space and details of the model being estimated. ML estimates on the population level a generic model space–linear models, neural networks–to extract information from data. Conversely, the inverse problem framework relies on a rigid model of physiological mechanics based on chemical or biochemical laws and processes to estimate unobserved model parameters. These estimated parameters are physiologically allowable values constrained by the structure of model equations and have inherent mechanical meanings, e.g., insulin clearance rate, because of a priori defined relationships within the model. Through the use of the mechanistic model, we both inject our a priori understanding of physiological mechanics and estimate unobserved parameters to characterize deep physiological knowledge [27][28]. As such, we change our ability to extract new physiological properties from data by projecting measurements onto model parameters within the inverse problem framework.

### Solving the inverse problem with data assimilation

The standard set of methods used to solve inverse problems is data assimilation (DA) [29]. Essentially, DA is a discipline and collection of computational methods that synchronize data with a mechanistic model, e.g., the hydrodynamics of the atmosphere for numerical weather prediction [30][31]. DA can also be interpreted as a Bayesian estimation procedure [32] that uses the model to map observations into posterior probability densities with either direct or variational methods [30][33][31][34]. DA has been used and developed in a long list of scientific and operational contexts ranging from landing on the moon [35] and aerospace in general [36], to robotics [37], but in medicine its application is just beginning [18][29].

Here we use DA within the inverse problem context in its smoother formulation to estimate model parameters, a process sometimes known as ‘calibration’ in statistics with computer models [38], using only the data of an individual patient. We smooth over a minimum time window length for which we have sufficient data to infer model parameters based on stochastic optimization using Markov chain Monte Carlo (MCMC) Method. Within Bayesian estimation procedures, we optimize model parameters by minimizing the prediction cost in glucose measurements in *L*_2_ norm with mean squared error (MSE). In this way, individual patient’s underlying physiological characteristics are encoded into personalized posterior distributions of MCMC-inferred parameter estimates.

The use of DA reverses a typical ML phenotype computation workflow by first personalizing a mechanistic model for an individual using only their data, producing effectively continuously-defined, time-specific, and individual phenotypes. This process generates continuous estimates of variables comprising states and parameters (e.g., blood glucose and insulin secretion) that function as new physiological biomarkers of the patient. The individual EHR sparsity issues are also managed by leveraging the chemistry/biochemistry constraints in model structure. The model, as a continuous-time representation of a system, handles data sparsity and missingness through synchronizing present individual data with the model using DA methods, and running the model at all other times. Specifically, the continuous-time model maps the transition of states and imputes model states with continuous simulation in time to fill in the gaps from missing data. In this way, the concept of missing data or irregularly spaced measurements is trivial.

### Our contribution

In this paper, we focus on extracting new phenotypes defined by unmeasurable biomarkers that quantify specific physiological properties, here insulin secretion, insulin clearance, and insulin action/resistance, that are not present in clinical observations. We developed and implemented a DA-driven methodological pipeline that computes continuous, high-fidelity phenotypes of properties not measured clinically to quantify patient physiological mechanics more deeply. The methodology shown in Figure 1 with a high-level structural overview on the left has three stages. We demonstrate the implementation by applying the pipeline to the glucose-insulin system for ICU patients over a specific time window, which can be generalized to other systems, as a use case to compute time-specific, deep physiological phenotypes.

**Figure 1:**
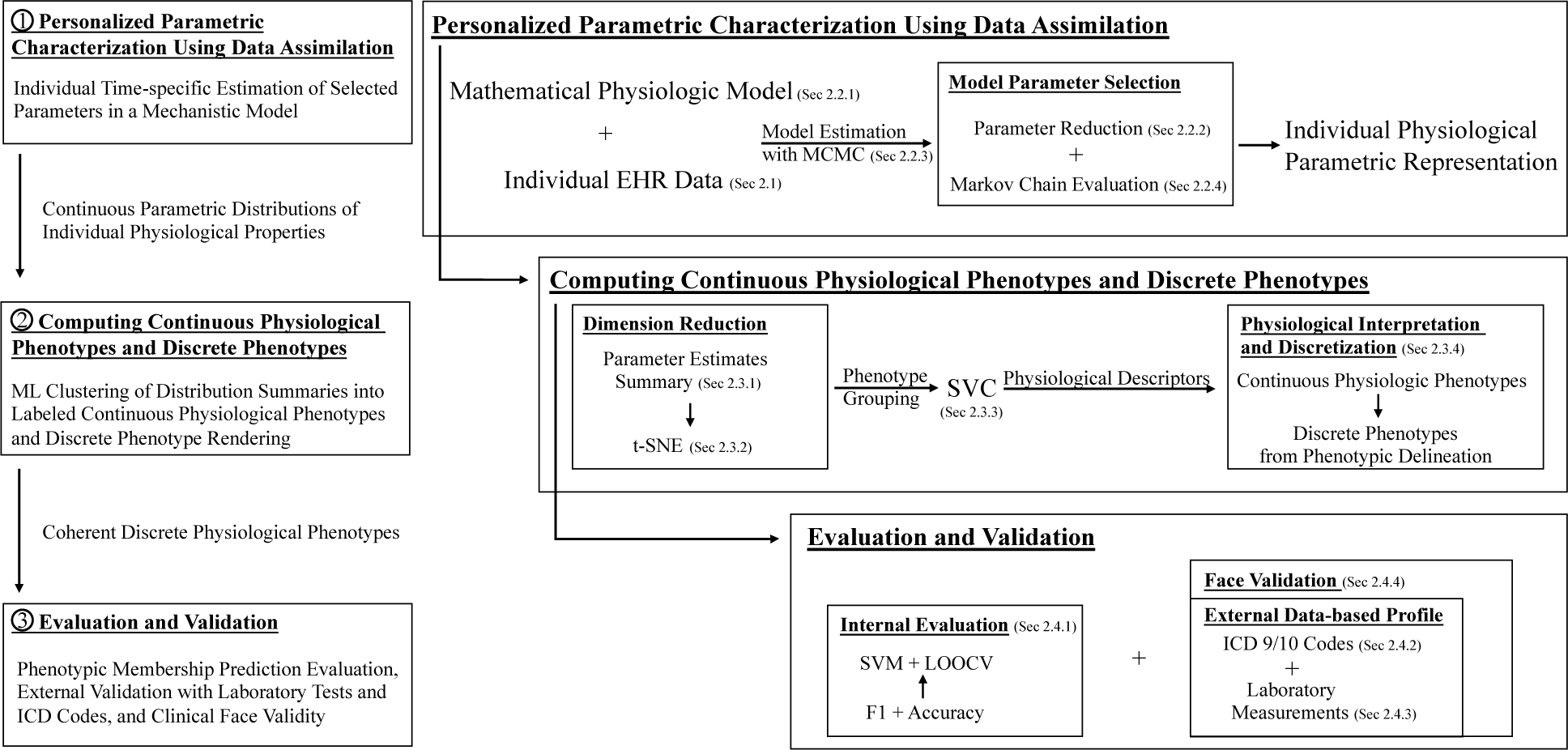
The methodological pipeline flow chart with a high-level structural overview on the left, delineated as three main stages: (i) personalized parametric characterization using data assimilation (DA), (ii) continuous phenotype computation with physiological interpretation using unsupervised ML and discrete phenotype rendering, and (iii) internal evaluation, analysis, and external validation of discrete phenotypes. The first stage computes personal continuous estimates from parametric characterization based on a mathematical mechanistic model using DA. The second stage clusters a cohort’s of individual estimates into phenotypic groups that have well-resolved physiological interpretations within each identified group. The third stage internally and externally validates the computer-derived phenotypes using laboratory measurements, ICD 9/10 codes, and face validity by an endocrinologist ICU expert.

The pipeline’s first stage estimates personal continuous distributions of measurable and unmeasurable physiological properties, including insulin secretion, insulin resistance, and insulin clearance, from personalizing the mechanistic ultradian model [22]–an a priori of the glucose-insulin system under known nutrition and exogenous insulin therapies. These estimates are computed by iterated MCMC samplings, creating a continuous parametric characterization of individual’s physiological mechanics that can be encoded into model parameter vectors. Tied to an individual patient, each vector is embedded with external physiological characteristics of glycemic functions provided by the ultradian model’s specificity.

The pipeline’s second stage computes phenotypes by clustering the convergent subset of physiological parameter vectors from the first stage into labeled groups. This stage implements unsupervised ML methods to partition the continuous parametric estimates of all patients into labeled continuous-space phenotypes imbued with physiological meanings. This stage also renders the discrete forms of the same phenotypes by delineating the continuous physiological phenotypes into discrete phenotypes.

While the quality of parametric estimates in the first stage is internally verified by minimizing the prediction MSE between model states and glucose measurements, the correctness of the parameterized physiological properties that are unmeasurable requires a new evaluative approach and is the pipeline’s third stage. This third stage evaluation begins by using the discrete phenotype rendering as proxy of extracted physiological properties for internal evaluation and external validation construction. Then, based on these discrete phenotypes and their qualitative clinical interpretations, this pipeline is evaluated quantitatively for predicting the phenotypic membership of each estimated ICU stay and for clinical coherency in three ways. First, we quantitatively summarize clinically observable differences using ICD 9/10 codes looking for coherency and consistency. Second, we quantitatively validate discrete phenotypes coherency and consistency against selected laboratory measurements, e.g., amylase, not used to create the phenotypes. Third, we externally validate qualitative discrete phenotypes through quantitative face validation by an endocrinologist to evaluate the practical clinical meaning and consistency. In this case, the endocrinologist examines the clinical mapping at each discretized physiological descriptor level based on explicit clinical notes and implicit health information that are associated with unmeasured insulin secretion, insulin resistance, and insulin clearance.

This methodology can be used to generate new information from ICU data that can plausibly scale to high-throughput, real-time settings, bringing new possibilities for optimal treatment strategies, computation, and future patient state prediction.

## 2 Methods

The three main stages in the methodological pipeline described above require data (Sec. 2.1) and comprise individual parametric characterization with a mechanistic model (Sec. 2.2), physiological phenotype computation (Sec. 2.3), and phenotype evaluation and validation (Sec. 2.4).

### 2.1 Data

#### 2.1.1 Data Set Construction

We use two ICU patient data sets in this work from two institutions, Columbia University and UCHealth Data Compass, both approved by the respective IRBs (Columbia University IRB AAAJ4503(M01Y06) and Colorado Multiple IRB 18-2519, respectively). To define experiment intervals in this paper for patient-level estimation from the EHR data originally identified by patient encounters, we identify two types of intervals: (i) ICU periods based on the recorded level of care, and (ii) regulated nutrition intervals based on the presence of a feeding tube with enteral nutrition (EN). Patient glycemic states estimation occurs during an ICU stay, defined by a contiguous ICU period longer than three days with EN. We also merge ICU stays when an enteral feeding tube is absent for less than 24 hours. For the remainder of this paper, we work with ICU stay ID as the identifier.

Table 1 details our two cohorts of ICU stays. We use cohort A from Columbia University (N=9) for model parameter selection (Sec. 2.2.2) and method configuration (Sec. 2.2.4) [39]. Cohort B (N=109) is used for phenotype computation (Sec. 2.3) and external validation (Sec. 2.4). We initially exclude pregnant patients from both cohorts. We identify cohort A by further excluding type 1 diabetes mellitus (T1DM) patients. To balance both the uncertainty in nutrition/insulin therapy and cohort breadth, the additional data inclusion criterion for cohort B is confining the ICU stay duration over 3 days with exogenous short-acting insulin only. Under this additional inclusion criteria, we identify cohort B (around 7% have T1DM) from a larger cohort with around 33.5% ICU stays (5157/15395) receiving exogenous insulin therapies.

**Table 1:** The nutrition and insulin therapy and demographic summary of ICU data sets cohort A, B, and B3 used in this work, including data resources, patients counts, ICU stays counts, data set inclusion criteria, ICU stay length in days, types of exogenous insulin therapy, insulin delivery counts, types of nutrition therapy, race, ethnicity, age, and gender. Neither CGM data nor insulin measurement was included, as is the general situation in an ICU setting. Cohort A comprises of nine manually-curated non-T1DM patients from the Columbia University Irving Medical Center Neurological Intensive Care Unit. Most patients required exogenous insulin therapies delivered through continuous intravenous infusions (mostly), bolus intravenous injections, and subcutaneous injections. Cohort B is a cohort of 109 patients receiving short-acting insulin under controlled EN with only brand-name nutrition recorded from the University of Colorado Health Data Compass Clinical Data Warehouse. Short-acting insulin was typically administered as a continuous intravenous infusion (5953 in total) and a limited portion is subcutaneous injection (6 in total). In each ICU stay, only measurements after the first non-zero nutrition were used to support this paper. From cohort B, cohort B3 comprises 45 ICU stays that have sufficient data in the last three-day period. Noting here that this table summarizes the complete ICU stays in cohort B3, not only for the last three-day period. A large portion of patients in cohort B3 was possibly on hemodialysis treatment due to acute kidney failure or chronic kidney disease. Glucose measurements and nutrition insulin therapy data were used to compute the model-based phenotypes from the pipeline. Other laboratory measurements and ICD 9/10 codes are used in external validation only.

| Data Summary Table |  |  |  |
| --- | --- | --- | --- |
| Summary | Cohort A | Cohort B | Cohort B3 |
| Data Resource | Manual Collection from Columbia University Irving Medical Center | Automatic Extraction from University of Colorado Health Data Compass Clinical Data Warehouse Between 2010–2019 | Same as Cohort B |
| # Patients | 9 | 109 | 45 |
| # ICU Stays | 9 | 109 | 45 |
| Inclusion Criteria | Non-pregnancy; Non-T1DM ; ICU Level of Care | Non-pregnancy; ICU Stay Duration in [3, 60] Days; Exogenous Insulin Only; Short-acting Insulin | Same as Cohort B |
| Stay Length (Days)<br>Mean±SD<br>Median [Min, Max] | 27±20<br>22 [13,78] | 8±6<br>6 [3,36] | 7±3<br>6 [3,19] |
| Exogenous Insulin | Continuous Intravenous Infusion; Bolus Intravenous Injection; Subcutaneous Injection (Probably Short-acting Only, But Uncertain) | Short-acting (Continuous Intravenous Infusion; Bolus Intravenous Injection; Subcutaneous Injection) | Same as Cohort B |
| # Insulin Delivery<br>Mean±SD<br>Median [Min, Max] | 39±36<br>30 [4,125] | 55±73<br>31 [1,387] | 104±87<br>75 [5,387] |
| Nutrition | TPN | TPN, No TPN | TPN, No TPN |
| Race |  | 83 White/Caucasian; 12 Black/African American; 4 Asian; 1 Multiple Race; 9 others | 37 White/Caucasian; 4 Black/African American; 1 Asian; 3 others |
| Ethnicity |  | 96 Non-Hispanic; 13 Hispanic | 41 Non-Hispanic; 4 Hispanic |
| Age<br>Mean±SD<br>Median [Min, Max] |  | 57±14<br>57 [25,90] | 64±13<br>65 [30,87] |
| Gender |  | 73 M; 36 F | 30 M; 15 F |

We use the following longitudinal data of the two cohorts from patient EHR records: administered TPN via continuous EN, non-bolus IV, and bolus IV deliveries; exogenous insulin therapies with continuous intravenous infusions, bolus intravenous injections, and subcutaneous injections; laboratory measurements synchronized with blood glucose measurement monitoring the general endocrine system state. Glucose measurements are more frequent (every 1-2 hours) when patients receive continuous infusion insulin therapies rather than subcutaneous injections. From these patient EHRs, we also use information about patient history to aid in validating computed phenotypes (Sec. 2.4.2 & Sec. 2.4.3).

#### 2.1.2 Time Window Partitioning to Balance Non-stationarity versus Data Sparsity

Patients in ICU are highly non-stationary with fast-changing physiological states that normally progress slowly, due to externally critical therapeutic interventions that affect internally cooperative human body mechanisms. We estimate patients only on a short time window to balance non-stationarity effects with data sparsity, conceptually managing a version of the variance-bias trade-off between temporal resolution and sufficient statistical representation. The time window for estimation is long enough to give robust statistical inference while managing non-stationarity, and the shortest possible with sufficient data required by individual-level modeling to produce an acceptably low error. For this reason, time window partitioning is a necessary consideration in the existing data sparsity case for individual-level modeling that requires joint data of glucose measurements, administration of insulin, and nutrition for one individual patient.

As such, we partition cohort B3 in the window of the last three-day period from cohort B, because this cohort not only satisfies the balance requirement but also gets the closest to stationarity with a well-defined aligning point of patients by enteral feeding tubes removal time based on either healthy or dying conditions. For completeness we also partition cohort B into two other sub-cohorts in the window of the first (cohort B1) and middle (cohort B2) three days that have different numbers of ICU stays than cohort B3 due to model estimation criteria and data availability.

### 2.2 Physiological Parameter Estimation to Characterize Individual ICU Stay

Here we will introduce the parameter estimation process within the DA smoothing framework to represent the physiological characteristics of individual ICU stay. This process personalizes a mathematical physiological model to characterize individual ICU stay as a physiological parameter vector in continuous distribution under qualification criteria.

#### 2.2.1 Mechanistic Model Representing Our Understanding of Physiological Functions

In this paper, we posit one model, the nonlinear mechanistic ultradian model [22], to represent our understanding of glucose-insulin physiological mechanics to limit the analysis for a robust solution. We use this model to inject personalized physiological hypothesis and extract mechanistic knowledge from observations to gain deep physiological insight. This relatively simple and widely validated model reflects essential features of the endocrine system and provides an unambiguous mathematized representation of the ultradian glycemic oscillation when patients are receiving enteral nutrition [40]. This model approximates the essential biological fidelity at a multi-day scale, which allows for estimation of physiological functions that are not directly measured or measurable but can sometimes be implicitly observed by clinicians; for example, the model estimation with an individual patient’s data can characterize insulin secretion. A schematic diagram of the ultradian model is shown in Figure 2, with all parameters interposed in all channels between the three main pools. A reader interested in technical details of the model mathematical formulas is referred to the Appendix (Sec. A.1).

**Figure 2:**
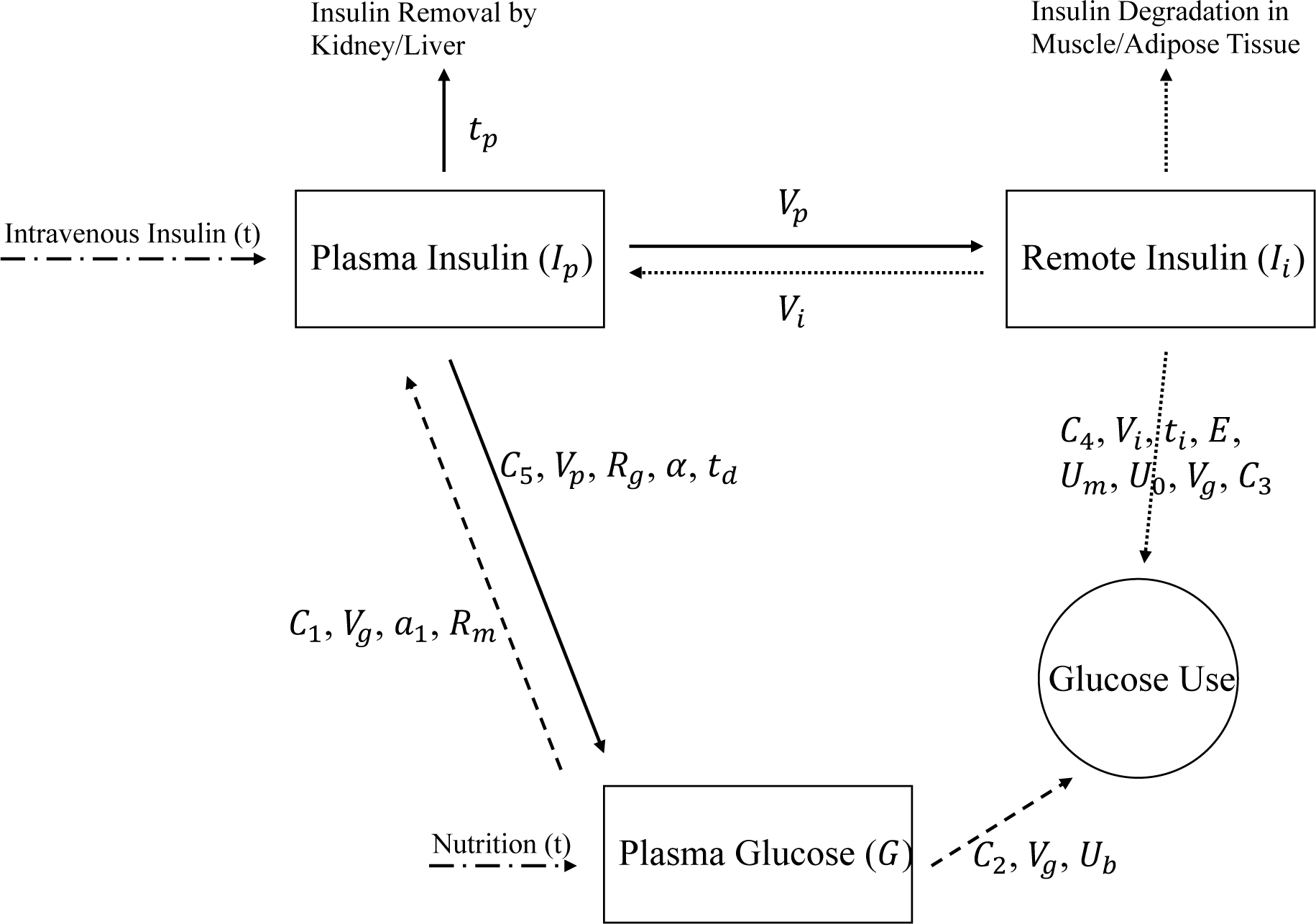
The ultradian model diagram. The glucose-insulin system consists of three main pools: the plasma insulin, the remote insulin in the intercellular space, and the plasma glucose. This non-autonomous ultradian model is represented by a system of ODEs with 6 states and 21 parameters. Exponential constants *a*_1_ and *C*_1_ affect insulin secretion; *t_p_* is the time constant for plasma insulin degradation by the kidney and liver; *R_g_* is the linear constant affecting insulin-dependent glucose production rate; *C*_3_ is the linear constant affecting remote insulin-dependent glucose removal rate in an implicit delayed process.

#### 2.2.2 Model Parameter Selection to Maximize Parameter Identifiability

Estimating the full suite of 21 parameters in the ultradian model [22] will cause identifiability issues, where data are insufficient to guarantee existence and uniqueness to infer solutions. Therefore, we further add a model selection procedure for model estimation to overcome the identifiability issues, which is identifying and estimating only a subset of parameters for model inversion. Fundamentally different from removing a set of parameters from the model to keep fewer elements, e.g., feature extraction with Lp regularization in a typical ML workflow, it is worth clearly pointing out that our approach keeps all parameters that govern the dynamics of the model. This model parameter selection procedure, at the expense of physiological fidelity, only selects which parameters we estimate and fixes excluded parameters with their nominal values when we estimate the model.

Specifically, we select model parameters from three perspectives: (i) the scalable rank-order optimization algorithm called parameter Houlihan method [41] that perturbs and picks the most influential parameters on glucose level (cf. Appendix A.2),(ii) the balance between model flexibility and fidelity to minimize forecast error with our non-clinical understanding of the mechanistic model structure, and (iii) clinically interest and potential utility.

Using cohort A, a triplet of parameters (*a*_1_, *t_p_*, *R_g_*) is adopted with the least MSE from nominees identified by the Houlihan method. This triplet is mathematically interpreted as a vector composed of exponential constant for insulin production inverse rate (*a*_1_), time constant for plasma insulin decay inverse rate (*t_p_*), and insulin-dependent glucose production rate (*R_g_*). Corresponding to the kidney-liver-pancreas subsystem, this parameter triplet is associated with pancreatic insulin secretion (*a*_1_), insulin clearance via liver and kidney (*t_p_*), and insulin resistance (*R_g_*), respectively. These endocrine physiological properties captured by the parameter triplet are important but implicitly quantifiable or unmeasurable/clinically unmeasurable. Because we select model parameters through transferable calibration that uses individual data, we adopt and estimate the same parameter triplet in cohort B for the rest of this paper.

#### 2.2.3 Markov Chain Monte Carlo (MCMC) Methods for Model Parameter Estimation

Given the mechanistic equations of the physiological model, we use Markov chain Monte Carlo (MCMC) with multiple Markov chains to estimate model parameters with uncertainty quantification (UQ) that reflects clinical reality, where each chain comprises the sampling iterations of MCMC parameter estimates. Herein we adopt a Metropolis-Hastings-within-Gibbs MCMC method, which is the Metropolis-Hastings algorithm incorporating a Gibbs sampler that updates parameter estimates one at a time [42]. This method, while less efficient than other MCMC methods, excels in cases where the full conditional probability cannot be determined analytically [43][44][45], which is the case when estimating a complex physiological model. For informatics researchers who are unfamiliar with the MCMC method, we note here that a Markov chain in the current context is a stochastic process of physiological model parameters in continuous state space, indexed by sampling time. We characterize each individual ICU stay as 10 Markov chains of model parameter estimates generated using different initial conditions, where each Markov chain is a sampled time series with 20K points.

#### 2.2.4 Markov Chain Evaluation of Estimation Accuracy and Convergence

The theoretically asymptotic properties and ergodicity [46] guaranteed by MCMC methods hinge on infinite computing resources. In practical implementation, the non-linear nature of MCMC without convexity prevents us from finding a unique solution as global error minima to represent model estimation, and Markov chains may fail to converge due to using a misspecified model [41][43]. It is prudent to screen chains for both prediction accuracy and convergence, especially the Metropolis-Hastings-within-Gibbs method is applied to estimate highly correlated parameters.

We construct a set of objective quality criteria to select from multiple Markov chains that have plausibly converged to global minima. Within multiple chains for a single ICU stay, we assess two types of chain convergence to determine inclusion: (i) intra-chain convergence of a single Markov chain to a stationary distribution, and (ii) cross-chain convergence to a common estimate. We access intra-chain convergence via Geweke statistic that compares the initial and terminal fractions of Markov chain iterations [47] with spectral density estimation to detect periodical behaviors, among all three parameters simultaneously. This is because that the beginning and the middle iterations in each Markov chain are abundant in variation of ‘drifts’ in nature as the chain typically starts somewhere far from the truth, while Geweke statistic identifies converged Markov chains that stay around equilibrium toward the end iterations without significant ‘drifts’. We explore complex across-chain convergence via the Gelman-Rubin (G-R) statistic that measures whether all Markov chains within a single ICU stay converge to a common solution, while setting chains burn-in–the beginning iterations to be thrown away–to be the first 80 percent of the total iterations for the statistic[48].

Markov chains independent by nature may converge to different solutions. To ensure parameter estimates represent the data, we choose for analysis all Markov chains both by intra-chain convergence in Geweke statistic and MSE close to the optimal MSE (+/- 2SD) of any chain. Chains significant in Geweke statistic (*α* = 0.06) are considered to have converged using the initial 3% and terminal 50% of iterates as Geweke fractions with technical details included in the Appendix (Sec. A.3). We also allow non-unique solutions for coexisting parameterized phenotypes within a single ICU stay, which were examined by G-R statistic for diagnosis purpose only.

### 2.3 Computing Continuous Physiological Phenotypes from Parameter Estimates and Rendering Discrete Phenotypes

In Sec. 2.2 we presented how individual physiological characteristics are encoded into continuous estimates of parameter vectors qualified by data-optimized MCMC sampling distributions to represent individual ICU stay. To compute phenotypes that represent cohort-scale categories from these estimates, we describe in this subsection a five-step process for grouping and labeling continuous-space physiological phenotypes, which we also delineate in discrete forms as discrete phenotypes for internal evaluation and external validation in Sec. 2.4.

#### 2.3.1 Summarizing Individual Parameter Estimates for Clustering

From each qualified MCMC chain, we construct an empirical distribution function (EDF) to represent physiological parameter estimates by retaining only the last 20% of sampling iterations with high probability region of the parameter space. Each EDF only keeps the variation around the equilibrium by excluding the variation of the ‘drifts’ in the full chain. This leads to each ICU stay to be associated with a dimension of (counts of selected Markov chains) * 4K iterations of each chain * 3 parameters.

To find shared phenotypic information among different patients, we then cluster a cohort of N patients into different groups from individual EDFs. The number of iterations in high dimensions, though, creates an unnecessarily large feature vector for clustering. For this reason, we summarize each individual EDF as a parametric 5D vector with five statistical indices–the position and the spread of the EDF–that can appropriately compress the function for each estimated parameter: mean, standard deviation (std), interquartile range (IQR), naive standard error of the mean, and the time-series standard error based on estimation of spectral density at 0. We pick these five indices because each parameter has its own encoded physiological properties and we want to comprehensively capture all properties–the ‘shape’–of different EDFs under every possible scenario. After this summary, each patient now has a dimension of (counts of selected chains) * 5 indexes * 3 parameters.

#### 2.3.2 Dimension Reduction of Parameter Estimates Summary

Dimension reduction is a common approach for improving clustering efficiency [49] and potentially configuring the number of groups in a cohort. Moreover, tuning hyper parameters for clustering methods is easier when we can visualize data points in high dimensions. The t-Distributed Stochastic Neighbor Embedding (t-SNE) method is a nonlinear unsupervised technique for projecting and visualizing high-dimensional data into a low-dimensional space [50][51] of 2D or 3D while preserving distributional features such as local similarity and global structure of the original space [51][52][53]. Compared with multidimensional scaling (MDS) methods, t-SNE does not involve eigen-analysis that MDS is usually based on. We use the t-SNE method before clustering patients to serve two purposes here: (i) mainly to further reduce the dimension of individual EDF summaries from 5D to 2D/3D, as the next clustering step maps points to an even higher dimension, and (ii) additionally to geometrically represent the cohort in 2D/3D based on an abstract notion of the distance between summaries. We select L-Infinity (Chebyshev) metric in t-SNE to distinguish scales in different parameter summaries. The t-SNE step reduces the feature dimension from (counts of selected chains) * 15D to (counts of selected chains) * 2D/3D t-SNE coordinate vector.

#### 2.3.3 Labeling Continuous-space Phenotypes by Clustering Common Characteristics

Following the t-SNE 2D projection, we use support vector clustering (SVC) for grouping stays and patients into labeled phenotypes comprised of continuously valued vectors of model-inferred physiological parameters. This unsupervised learning kernel algorithm identifies regions of nearby t-SNE coordinates–possibly belonging to different patients–most efficiently bounded by a set of small circles in the same 2D space through feature mapping in high dimensions [54]. This feature identification is implemented numerically in Matlab [55][56] using the statistical pattern recognition toolbox (stprtool) [57]. For SVC methodological choices, we choose the radial basis function (RBF) kernel and adjust the smoothness and the fraction of outliers allowed in a cluster for coefficients iteratively until visually sensible. Each SVC-identified cluster determines a labeled continuous-space phenotype with parametric representation of underlying physiological mechanics.

A patient whose t-SNE coordinates appear in multiple SVC clusters may be associated with multiple phenotypes. This happens because (i) an ICU stay can pertain to coexisting phenotypes with shared physiological features due to, e.g., highly non-stationary insulin clearance at early stage caused by critical illness treatments, and (ii) our method allows coexisting phenotypes as cross-chain convergence of independent Markov chains are not strictly examined for inclusion. For a better phenotypic interpretation at the ICU-stay/patient level, though, we further identify coherent clusters as those containing t-SNE coordinates from the same ICU stay.

#### 2.3.4 Extracting Physiological Definitions and Delineating Discrete Forms

We extract the physiological meanings of SVC-identified clusters–without prior information of phenotypic grouping–both on the continuous and discretized parameter space using the following four steps: (i) generating phenotypic-level parameter estimates in continuous forms by concatenating the empirical distribution functions (EDFs) within each SVC-identified cluster, (ii) extracting physiological meanings of continuous-space phenotypes using the unique physiological descriptors of the parameter triplet (*a*_1_, *t_p_*, *R_g_*) from the ultradian model, (iii) producing coherent continuous phenotypes by expanding SVC-labeled clusters with auxiliary groups–ICU stays associated with multiple SVC clusters–and dropping small-sample groups, and (iv) delineating discretized phenotypic patterns using a customized ranking relative to the average patient glycemic states in ICU. After these four steps, each discrete phenotype with extracted physiological interpretation is potentially viable for clinical mapping, which is implemented in an evaluation and validation process in the next section. We only analyze the phenotypes computed in the last three-day period (cohort B3), although we also compute phenotypes in the first (cohort B1) and middle three days period (cohort B2) as well for completeness and to show readers what phenotypes exist at different time periods.

Our main phenotyping results are labeled continuous phenotypes that are coherent with physiological interpretations defined in our third methodological step. However, to make evaluation and validation efforts that comprise the remaining sections clearer, we focus on the discrete forms of these phenotypes. We refer to the discrete forms of these phenotypes simply as discrete phenotypes hereafter unless specified.

### 2.4 Evaluating and Validating Discrete Physiological Phenotypes Against Laboratory Measurements, ICD 9/10 Data, and Clinical Notes

Model-derived phenotypes are potentially clinically valuable to provide clinical decision support when they can be mapped to practical clinical meanings and validated. We verify the model’s ability to represent blood glucose measurements data for inversion estimation in the metric of MSE. Similarly, we need to validate model-derived phenotypes defined by parameter estimates of physiological functions are not random and non-sensible, but accurate with potentially clinically meaningful structure. However, a lack of direct measurements of the same physiological functions hinders the construction of direct validation of these model-inferred physiological properties. Specifically, there are currently no direct measurements of insulin secretion, insulin clearance, and insulin action/resistance. Nevertheless, we can use recorded patient data that are clinically associated with these functions as an external validation of our model-derived phenotypes. As such, we evaluate and externally validate discrete phenotypes as proxies of model-inferred physiological functions in the space of phenotypic labels, laboratory measurements, ICD 9/10 codes, and clinician’s proxies of the same physiological functions: (i) quantitative evaluation of phenotypes reliability to predict the phenotypic membership of each estimated ICU stay, (ii) quantitative validation against external data from ICD 9/10 codes, (iii) quantitative validation against external data from selected laboratory measurements, and (iv) quantitative chart review with face validation by an endocrinologist and expert in ICU.

#### 2.4.1 Statistical Evaluation of Phenotype Labeling by Predictive Power

To quantitatively evaluate the reliability of model-derived phenotypes, we use discrete phenotypic information to predict the phenotypic membership of each estimated ICU stay. We access the predictive power of phenotype computation via F1-score and average accuracy using the support vector machine (SVM). This algorithm classifies features of each EDF labeled with identified phenotypes. We use leave-one-out-cross-validation (LOOCV) to tune SVM-required configuration for robust prediction, where the tuned SVM parameters reach the highest average accuracy for multi-class labels prediction.

#### 2.4.2 Summarizing Phenotypic Difference Using ICD 9/10 Codes

To find evidence of coherence and consistency, we analyze the correspondence between phenotypic differences in physiological interpretations and clinical observable differences. Specifically, we count the occurrence of selected ICD 9/10 codes distributed to patients in each phenotype and then check inter-phenotype distinctiveness based on the difference in percentage counts.

#### 2.4.3 External Validation Against Remaining Laboratory Measurements

We statistically test each phenotype’s coherence and consistency with external laboratory measurements. From EHR data we analyze laboratory tests that are associated with organ functions encoded in parameter parameters and excluded from phenotype computation. We then use a Welch two-sample t-test to test the mean value difference of patients’ laboratory measurements in each phenotype to conclude clinical coherency and consistency.

#### 2.4.4 Clinical Face Validity of Discrete Phenotypes at Physiological Descriptor Level

To further externally validate that the model-derived phenotypes have correctly captured physiological characteristics from well-resolved ICU stays, we use face validation [58] to confirm whether the discrete phenotypes accurately represent the patients from an endocrinologist’s opinion based on a quantitative chart review of patient charts. Face validation is a sanity check based on experts’ opinions that provides a crude confidence quantification of the sensibility and relevance of the modeling results, including evaluation of methods and conclusions. We quantitatively check the computer-to-clinical mapping with face validity at the discretized physiological descriptor level: the estimated parameter triplet (*a*_1_, *t_p_*, *R_g_*) with their associated physiological interpretations (cf. Sec. 2.3.4). This mapping externally validates model-inferred physiological functions that are not or cannot be directly measured, but can be inferred indirectly by clinicians. For each patient, we ask the clinician through a blind face validation to categorize each of the three physiological descriptors into three bins from low, medium, and high. Recall that phenotypes are associated with the modeled kidney-liver-pancreas subsystem through the unmeasured parameter triplet: insulin secretion (*a*_1_), insulin clearance (*t_p_*), and insulin resistance (*R_g_*).

## 3 Results

### 3.1 Physiological Model Parameter Estimation to Characterize Individual ICU Stay

Within the DA smoothing framework, we generated Markov chains of model estimates and selected chains under qualification criteria to parametrically represent individual physiological characteristics. We estimated each ICU stay using 10 Markov chains of the parameter triplet (*a*_1_, *t_p_*, *R_g_*), where each chain was a sampled time series with 20K points of estimates for each parameter. At this stage, the dimension was 20K * 3 parameters = 60K for each Markov chain and 10 * 20K * 3 = 600K in total for each patient. Then we used Geweke statistic and MSE to select qualified Markov chains. Figure 3 illustrates the Markov chains selection (cf. Sec. 2.2.4) applied to one ICU stay from cohort A as an example. There was consistent MSE among the chains, particularly for those with low MSE. Also, given the narrow bandwidth, every chain had at least one convergent parameter (indicated in blue) based on fraction-identified Geweke statistics. For this ICU stay, we selected the seven chains indicated in black outline.

**Figure 3:**
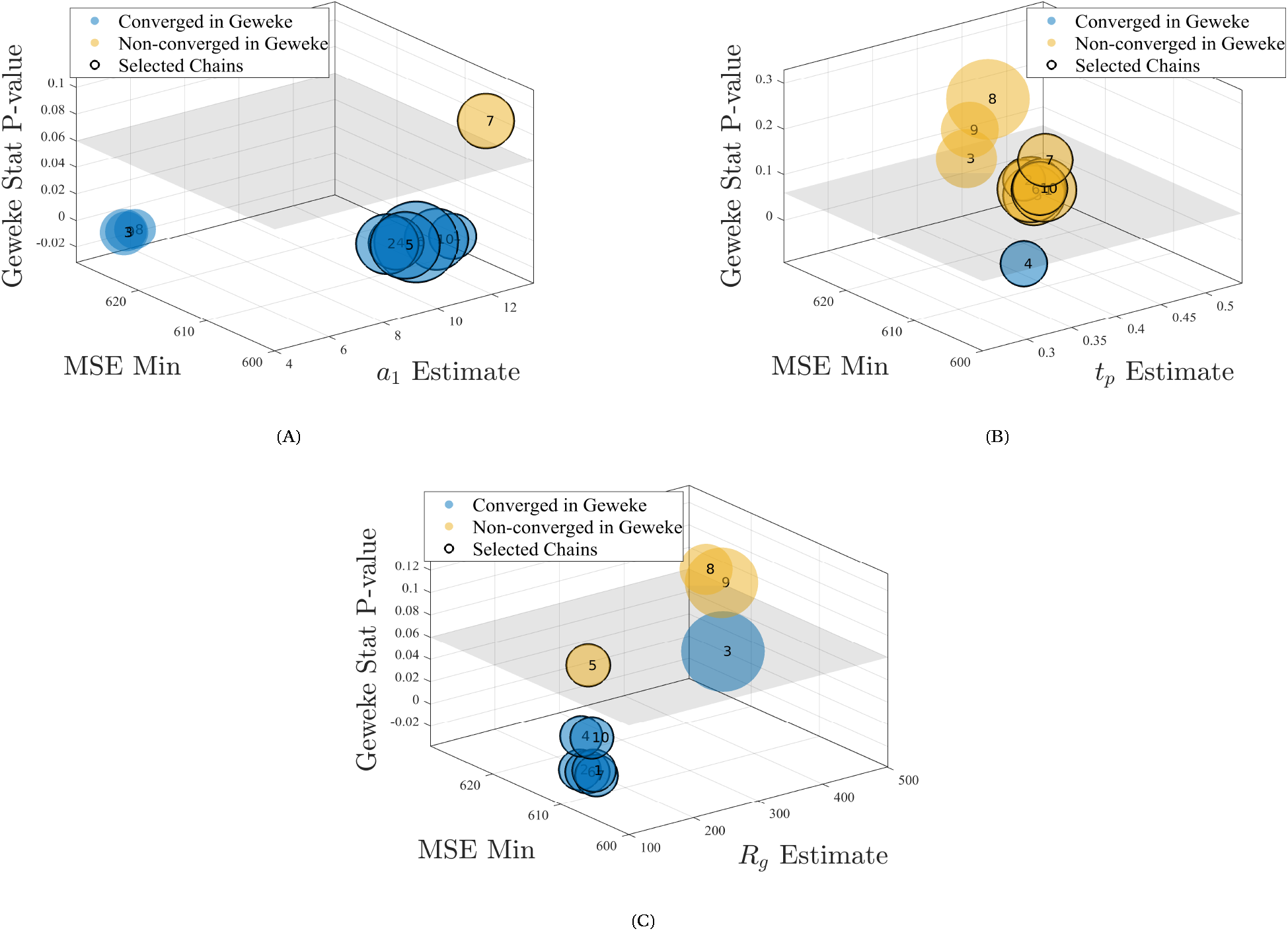
Markov chains selection to parametrically characterize a patient. Colored bubble charts of Markov chains selection for all 10 chains of one ICU stay from cohort A, with axes of parameter estimate, MSE, and Geweke statistics p-value, for each model parameter **(A)** *a*_1_, **(B)** *t_p_*, and **(C)** *R_g_* separately. Each bubble representing a single Markov chain was labeled with chain number. Bubbles were drawn with radius corresponding to Markov chain IQR and colored blue to indicate Geweke convergence of that parameter. Markov chains with low MSE and intra-chain convergence of at least one parameter were selected. The seven Markov chains meeting these criteria were outlined in black and were selected to represent this ICU stay.

### 3.2 Computing Continuous Physiological Phenotypes from Parameter Estimates and Rendering Discrete Phenotypes

Note here that all previous steps in the results were demonstrated using the smaller cohort A. Starting from this section unless otherwise noted, all steps in the pipeline were conducted on cohort B3 to better illustrate the phenotype computation and validation process.

#### 3.2.1 Summarizing Individual Parameter Estimates as Statistical Indexes

Based on parametric representation of each individual ICU stay, parameter estimates of all ICU stays in cohort B3 were represented by 330 EDFs of qualified Markov chains, where each EDF was represented by the last 20% sampling iterations of a selected chain. To facilitate phenotype labeling from segmenting these estimates using clustering methods, we first compressed each EDF into five statistical indexes. In this summary step, we transformed 330 EDFs representing cohort B3 into 330 vectors in 5D, which also reduced the total dimension from 330 * 4K * 3 = 3960K to 330 * 5 * 3 = 4950.

#### 3.2.2 Dimension Reduction of Estimates Summary for Cohort Segmentation

Using t-SNE, we reduced the dimensionality of 330 5D vectors into 2D/3D coordinates while leaving the total counts of vectors unchanged (cf. Sec. 2.3.2). We manually tuned the t-SNE perplexity parameter as 30 from the normal range (10,50) for balancing local embedding with global structure. Although t-SNE in 3D could potentially help reduce cluster intersections, we chose 2D over 3D that further reduced the total dimension from 330 * 5 * 3 = 4950 to 330 * 2D = 660 as 330 t-SNE 2D vectors. This is because t-SNE 2D presentation given our properly tuned parameters already shows clear separation in the cohort with a better visualization. Figure 4 (inset) shows the raw, un-clustered t-SNE result with a clear structural separation in the lower left and top left area, indicating at least three possible phenotypes in cohort B3. Noting here that the t-SNE 2D axes are arbitrary and meaningless.

**Figure 4:**
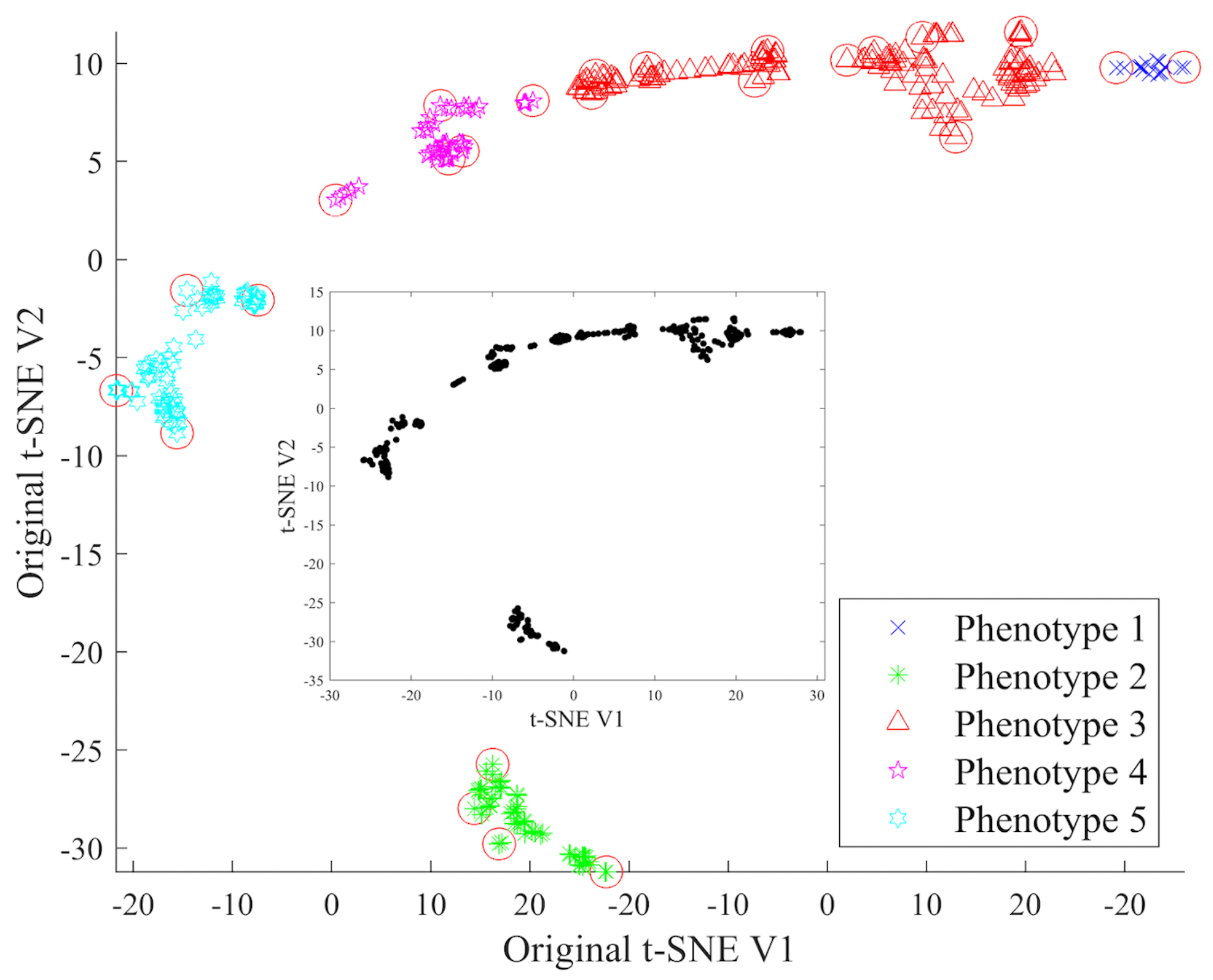
**Step 1 (inset)**: Raw and un-clustered t-SNE dimension reduction of selected chains summary of 45 ICU stays in cohort B3 with specified methodological choices. **Step 2**: Colored phenotypic label computation with SVC that clusters t-SNE coordinates of chains summaries based on SVC methodological choices. Circled coordinates are support vectors mapped to the surface of the high dimensional feature space sphere. Five clusters were identified indicating five potential phenotypes.

#### 3.2.3 Identifying Continuous-space Phenotype Labels within the Cohort

In order to compute cohort-level labels of continuous phenotypes, we used SVC to group parametric representations of ICU stays based on nearby t-SNE 2D coordinates (cf. Sec. 2.3.3) without changing the dimension. As a potent internal validation, this process was clustering ICU stays without knowing how patients were partitioned in the EHR data. Figure 4 shows five SVC-identified labeled phenotypes in cohort B3 using a RBF kernel radius of 4 and boundary penalization parameter of 0.6, indicating five potential phenotypes with each phenotype having a unique centroid in the 2D space. A similar process yielded three and four clusters for cohort B1 and B2, respectively.

#### 3.2.4 Extracting Physiological Definitions of Labeled Continuous Phenotypes and Rendering Discrete Phenotypes

We reconstructed physiological information from the SVC-identified clusters in four steps to map the labeled physiological phenotypes to clinical interpretations (cf. Sec. 2.3.4). Based on two steps that extracted the physiological meanings of labeled continuous-space phenotypes using physiological descriptors of triplet (*a*_1_, *t_p_*, *R_g_*), Figure 5 shows the continuous parametric representations of five distinct phenotypes identified from SVC in cohort B3 in 2D and 3D median space. These five distinctive densities of parametric phenotypes revealed five unique physiological traits. Recall that these phenotypes were associated with unmeasured insulin secretion (*a*_1_), insulin clearance (*t_p_*), and insulin resistance (*R_g_*).

**Figure 5:**
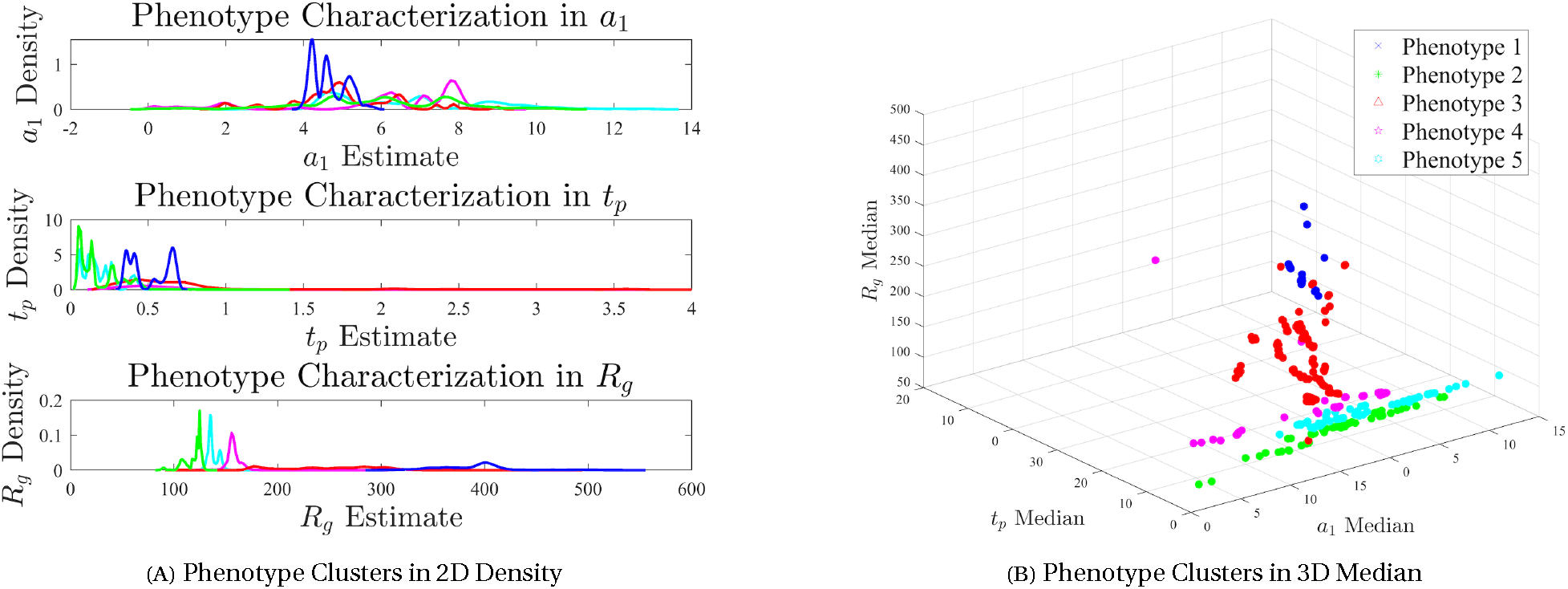
Continuous parametric characterization of labeled phenotypes from SVC in cohort B3 with density plots in **(A)** 2D and **(B)** 3D median space. Clusters coloring was the same as Figure 4. Both density plots in 2D and 3D median space showed five different distribution patterns over the entire parametric space and thus revealed five different phenotypes. For example, the phenotype in blue was interpreted as the phenotype with patients that had *a*_1_, the exponential constant for insulin secretion inverse rate, roughly between 4 and 5; *t_p_*, the time constant for plasma insulin clearance inverse rate by the kidney and liver, between 0.25 and 0.7 min; and *R_g_*, the insulin-dependent glucose production rate or insulin resistance, between 300 and 550 mg/min.

Thirdly, we produced coherent phenotypes from SVC-identified clusters. For example, we previously identified five distinct phenotypes from SVC clusters in cohort B3. We augmented these five phenotypes with five auxiliary phenotypes that were associated with multiple SVC clusters at the ICU-stay level. Then five out of these ten phenotypes were associated with fewer than three ICU stays and we dropped them considering statistical power. We ended with five coherent, labeled continuous phenotypes that had well-resolved ICU stays, meaning that each phenotype had patients with uniform physiological functions from parameterization.

In the fourth step, we generated discrete phenotypes by delineating phenotypic patterns using a qualitative customized ranking relative to the average glycemic states in ICU. Table 2 tabulates the delineation of coherent discrete phenotypes along with discretized physiological qualities identified in cohort B1, B2, and B3. We also used this table to show that phenotypes identified in the first three days were likely not to show up in the last three days, as patient health status changed drastically during the entire ICU stay. Similar ‘relative’ phenotypes, e.g., G7 and G9, were not the same phenotypes as they were computed from different periods. Nevertheless, observation of the ‘same’ phenotypes across three periods could indicate that patients’ sickness condition was not improved. More complete technical descriptions of the ranking method are provided in the Appendix (Sec. A.4).

**Table 2:**
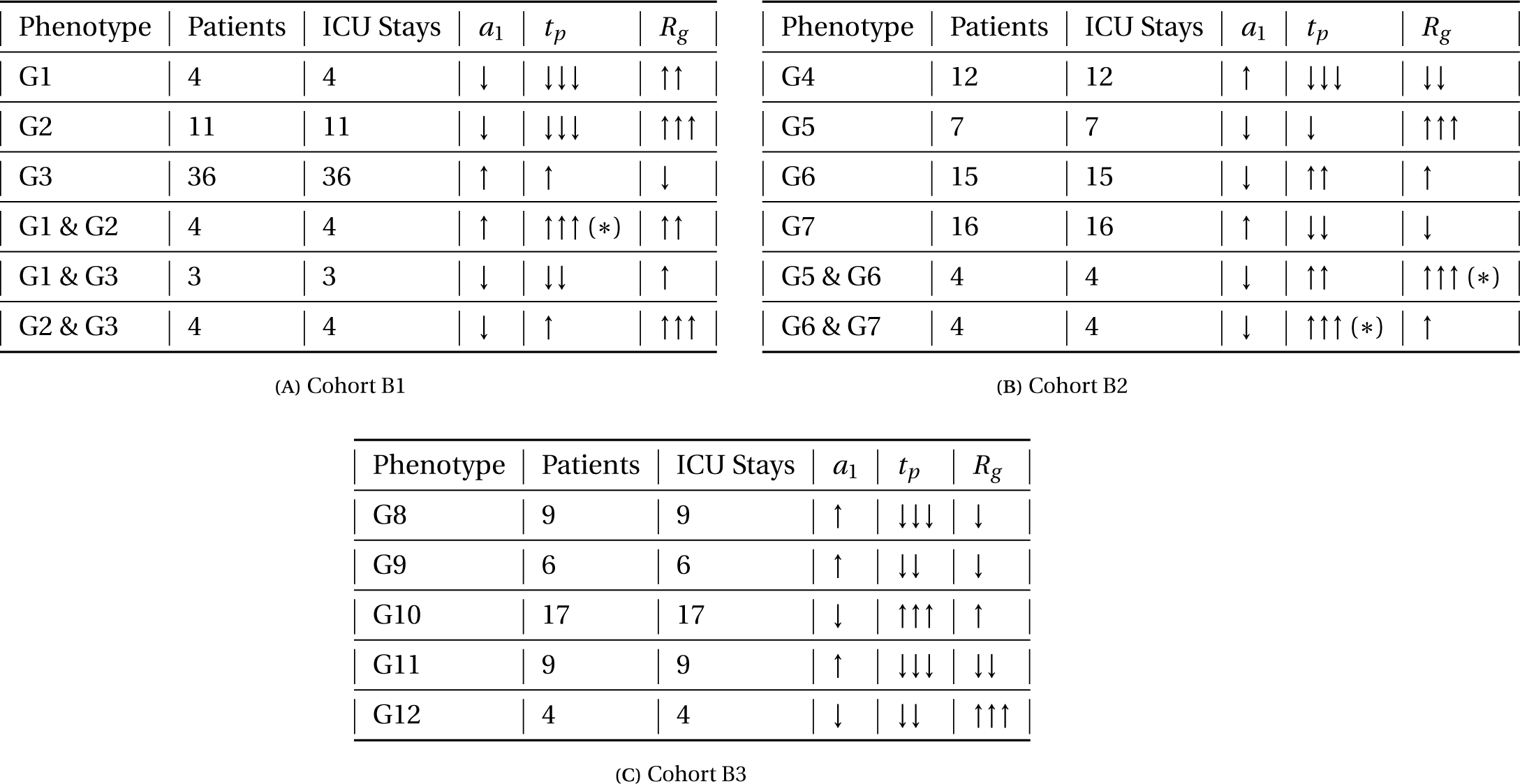
Phenotype delineation of coherent discrete phenotypes identified in cohort **(A)** B1, **(B)** B2, and **(C)** B3. Five phenotypes were identified in cohort B3, six in B1, and six in B2. Table columns correspond to phenotypes, number of patients, number of ICU stays in each phenotype, and parameter ranking of the parameter triplet (*a*_1_, *t_p_*, *R_g_*): *a*_1_, the exponential constant for insulin secretion inverse rate; *t_p_*, the time constant for plasma insulin clearance inverse rate; and *R_g_*, the insulin resistance. Within each cohort, for each phenotype the customized ranking method compared parameter triplet estimation average to the posterior average across all phenotypes–the midpoint of all parameter estimates. We qualitatively discretized estimation into six bins centered at the midpoint. The number of upward and downward arrows indicated a ranking of how far from this midpoint each phenotype average was, ranging from 0 to double the midpoint value. Phenotype average that fell outside of either end of the range had a complementary star sign (*). For example, phenotype G8, identified in cohort B3, was interpreted as having patients with insulin secretion rate relatively slightly below the mean, insulin clearance rate much higher than average, and slightly decreased insulin resistance. We named auxiliary phenotypes as the incorporation of SVC clusters names they were previously associated with and then separated from. For example, auxiliary phenotype ‘G1 & G2’ in cohort B1 was separated from previously SVC-identified clusters G1 and G2.

Note that the third and fourth step outputs were simply two renderings of the same phenotype computation. While we used the discrete phenotypes–the fourth step outputs–for evaluation and validation with clinical interpretations, our main phenotyping results were the labeled continuous phenotypes that were coherent and corresponded to specific physiological interpretations produced in the third step of our pipeline.

#### 3.2.5 Discrete Phenotype Contextual Interpretation with Laboratory Measurements and ICD 9/10 Codes

To potentially guide clinical ontology, we incorporated physiological interpretations of time-specific discrete phenotypes with data-based profiles of patients to visualize clinically reviewable patient progression. The selected laboratory measurements and ICD 9/10 codes in data-based profiles best reflect organ functions of the liver, pancreas, and kidney that are captured by estimated parameters (cf. Appendix A.5). As one possible hierarchical visualization based on organ functionalities, Figure 6 starts with five discrete phenotypes G8-G12 identified in cohort B3 on the left hierarchy steps, and progresses to synthesized time-specific descriptions with clinical characterizations on the right. This delineation process demonstrates how the hierarchical visualization based on synthesized phenotypic picture can be potentially helpful to differentiate patients in a clinical context, using new biomarkers defined by discretized estimates of physiological functions. There may be clinically relevant ways to interpret the phenotypes with different delineation steps and characterize clinical properties associated with them.

**Figure 6:**
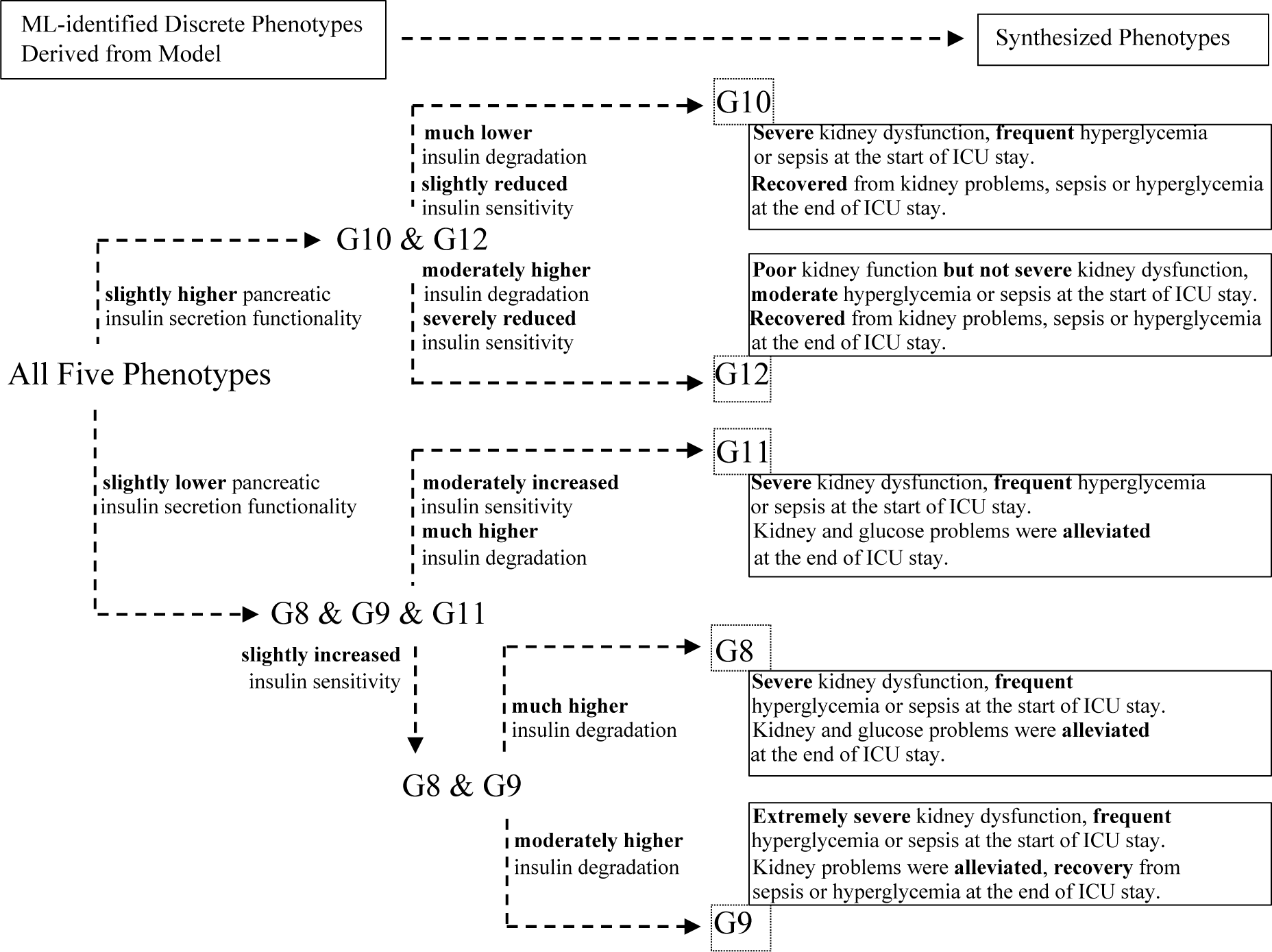
Discrete phenotype contextual interpretation with corresponding synthesized description based on model-inferred phenotypic delineation. This figure is a hierarchical visualization of patient progression by delineating time-specific discrete phenotypes identified in cohort B3. This visualization is only one example of contextual interpretation in a clinically relevant way. Starting from left, we separated discrete phenotypes G8-G12 at each hierarchy based on model-inferred organ functionalities, and ended with the synthesized interpretations combining clinical observations on the right. Comparing the level of pancreatic insulin secretion functionality at the first hierarchy, phenotypes G8, G9, and G11 could be differentiated from G10 and G12. Then comparing the level of insulin clearance at the second hierarchy, G10 could be differentiated from G12. Similarly, comparing the level of insulin resistance at the second hierarchy, G11 could be differentiated from G8 and G9, followed by comparing the level of insulin clearance at the third hierarchy to differentiate G8 from G9.

### 3.3 Evaluating and Validating Discrete Physiological Phenotypes Against Laboratory Measurements, ICD 9/10 Data, and Clinical Notes

To establish the clinical context of the model-derived physiological phenotypes, in this section we focused on those five discrete phenotypes G8-G12 identified in cohort B3–the last three-day period of cohort B–for reasons detailed in Sec. 2.1.2. As a reminder, these five phenotypes toward validation were simply rendered from the same computation process that also rendered continuous physiological phenotypes–the main phenotyping results in this paper. Because there is a lack of direct measurements of model-inferred properties, we externally validated the discrete forms of these five phenotypes against EHR data of laboratory measurements, ICD 9/10 codes, and clinical interpretations. Note that these data were not used previously in the model-based phenotyping pipeline and because of this, we can use this contextualization as a validation of the model-derived phenotypes.

#### 3.3.1 Quantitative Evaluation of Predicting Phenotypic Membership of Parameterized ICU Stays

As we used SVC to group parametric representations of ICU stays into clusters of t-SNE coordinates (cf. Sec. 3.2.3), we chose to use support vector machine (SVM), which shares a similar mechanism with SVC, to quantitatively access phenotypes predictive power. We picked a radial kernel for SVM and tuned kernel parameters with LOOCV to classify summary statistical features of 330 EDFs labeled with phenotypes G8-G12 (cf. Sec. 3.2.4). The prediction macro F1-score was 88.3%, the weighted F1-score was 89.3%, and the average accuracy was 89.4%, showing that the ML labeling of phenotypes was statistically predictive.

#### 3.3.2 Quantitative Summary of Clinical Observable Difference Using ICD 9/10 Codes

To find clinical evidence that supports difference between phenotypes, we calculated and compared percentage counts of eight selected ICD 9/10 codes (cf. Appendix A.5) for patients in each discrete phenotype. The observable physiological differences with regard to organ functionalities revealed by these ICD 9/10 codes were in general consistent with the findings of five discrete phenotypes, which also supported the contextual interpretation illustrated in the hierarchical visualization in Sec. 3.2.5.

#### 3.3.3 Quantitative External Validation Using Laboratory Measurements Excluded From Phenotype Computation

To validate the clinical coherence and consistency of model-inferred physiological functions against observed patient physiological states in laboratory measurements excluded from phenotype computation, we analyzed the data-based profiles based on five selected laboratory measurements (cf. Appendix A.5) of patients in each discrete phenotype. Table 3 contains a statistical summary of patients laboratory values for phenotypes G8-12, for the period of the first and last three days, respectively. Through this table we found evidence of the effective separation between phenotypes (horizontal comparison). For example, in the last three-day period, we found high counts of creatinine (CREAT) test for phenotype G10, which relates to kidney functionality in the absence of dialysis treatments. We also observed frequent lipase (LIPA) tests only for phenotype G11 in the last three-day period, which suggests clinical concern for pancreatic functions in this group. In the table were also records of elevated levels of aspartate aminotransferase (AST) tests indicating potential hepatic issues for phenotypes G8 and G10. Additionally, this table also shows that patient’s health status changed consistently over time in the same phenotype (laboratory-wise vertical comparison). Besides the summary presented in Table 3, we also statistically tested selected laboratory mean values among all phenotypes with a Welch two-sample t-test to check inter-phenotype clinical differences, which was synthesized into the phenotypic description in the next section.

**Table 3:** Data-based profiles of five selected laboratory tests (cf. Appendix A.5) measured in patients from phenotypes G8-G12 in the last three-day period, and in the first three days of the same patients. Table last five columns corresponded to phenotypes with their patients counts, five rows corresponded to selected laboratory tests, and each sub-row consisted of measurements summary within the laboratory test for the specific period, including measurements counts, mean*±*SD, and median [min, max]. Quantities not measured for each phenotype were noted as NA. Interestingly, laboratory measurements related to liver/kidney function, including alanine aminotransferase, albumin, and bilirubin, were not present in any phenotype identified in the last three days, such that they were not included into the data-based profiles of the first three-day period.

| Data-based Profiles with Laboratory Measurements |  |  |  |  |  |  |  |
| --- | --- | --- | --- | --- | --- | --- | --- |
| Laboratory (Last Three & First Three Days) |  |  | G8(N=9) | G9(N=6) | G10(N=17) | G11(N=9) | G12(N=4) |
| AMY | # Measurements<br>Mean±SD<br>Median [Min, Max] | Last | 1<br>15.0 | NA | 3<br>15.7±3.1<br>15.0 [13.0,19.0] | 1<br>39.0 | NA |
|  |  | First | NA | 1<br>22 | 3<br>15.3±2.5<br>15.0 [13.0,18.0] | NA | NA |
| AST | # Measurements<br>Mean±SD<br>Median [Min, Max] | Last | 27<br>698.9±908.6<br>446.0 [10.0,4240.0] | 10<br>33.6±17.8<br>27.0 [19.0,75.0] | 27<br>1262.0±3652.0<br>47.0 [20.0,15920.0] | 12<br>148.8±155.9<br>73.5 [25.0,415.0] | 6<br>39.2±24.2<br>25.5 [21.0,78.0] |
|  |  | First | 28<br>1118.5±1160.3<br>815.5 [15.0,4240.0] | 8<br>89.8±141.1<br>21.0 [12.0,420.0] | 32<br>1076.0±3373.0<br>47.0 [22.0,15920.0] | 16<br>117.1±121.1<br>84.5 [13.0,415.0] | 5<br>41.8±26.0<br>25.0 [21.0,78.0] |
| LIPA | # Measurements<br>Mean±SD<br>Median [Min, Max] | Last | NA | NA | NA | 16<br>66.5±76.9<br>44.0 [23.0,338.0] | NA |
|  |  | First | 1<br>9 | 1<br>15 | NA | 19<br>77.8±110.5<br>17.0 [10.0,338.0] | NA |
| BUN | # Measurements<br>Mean±SD<br>Median [Min, Max] | Last | NA | 2<br>51.5±17.7<br>51.5 [39.0,64.0] | 3<br>16.3±11.4<br>13.0 [7.0,29.0] | 1<br>33 | 1<br>40 |
|  |  | First | 3<br>21.0±7.0<br>18.0 [16.0,29.0] | 4<br>30.3±11.1<br>27.5 [20.0,46.0] | 6<br>27.8±12.9<br>26.5 [13.0,47.0] | 5<br>17.0±11.7<br>17.0 [6.0,36.0] | 3<br>29.7±17.7<br>21.0 [18.0,50.0] |
| CREAT | # Measurements<br>Mean±SD<br>Median [Min, Max] | Last | 79<br>2.3±1.1<br>2.4 [0.7,4.4] | 50<br>2.2±1.0<br>1.9 [1.2,4.3] | 119<br>1.1±0.5<br>1.0 [0.3,3.4] | 58<br>1.3±0.8<br>1.1 [0.5,3.8] | 23<br>2.3±0.8<br>2.6 [0.6,3.0] |
|  |  | First | 81<br>1.8±0.8<br>1.4 [0.8,4.0] | 53<br>1.6±0.8<br>1.4 [1.0,4.3] | 125<br>1.1±0.4<br>1.1 [0.6,2.7] | 69<br>1.3±0.8<br>1.2 [0.5,3.8] | 23<br>2.3±0.8<br>2.6 [0.6,3.0] |

#### 3.3.4 Synthesizing Qualitative Phenotypic Description Using Laboratory Measurements and ICD 9/10 Codes

To summarize phenotype coherence from a clinical perspective, we generated qualitative descriptions by synthesizing physiological interpretations with external data analysis in previous two sections. Table 4 uses phenotype G8 as an example to illustrate the phenotypic synthesis in cohort B3. The synthesis process also defined a map from the unmeasurable parameterized phenotypes to the measurable data-based profiles, which aided in the analysis and construction below.

**Table 4:** Steps in generating synthesized descriptions for phenotype G8 in cohort B3. We constructed a comparative baseline from the data of ICU patients given only short-acting insulin therapy. This baseline represents the average patient glycemic states in a typical ICU environment, where critically ill patients primarily receive intravenous infusion of insulin. We summarized the concordance and discordance between model-based phenotypes and data-based profiles, organically combining the physiological interpretations. For each ICD 9/10 code, the percentage count was dividing the code counts in each phenotype by the total number of distinct ICU stays, respectively. Missing laboratory measurements were noted. Also noting here that cohort B3 reported no obese patients and few with diabetes per ICD 9/10 codes.

| Qualitative Interpretations of Phenotype G8 |  |
| --- | --- |
| Steps | Descriptions |
| Step 1:<br>Model-based Phenotypic Interpretation | Patients in phenotype G8, compared to our baseline, have: a slightly decreased pancreatic insulin secretion, much higher insulin clearance assuming no dialysis, and a slightly decreased insulin resistance. This interpretation is based on extracted physiological definitions indicating the slightly higher (↑) insulin secretion rate, much lower (↓↓↓) insulin clearance rate, and slightly lower (↓) insulin-dependent glucose production rate. Compared to the nominal model parameter values, patients in G8 have a lower pancreatic insulin secretion functionality, higher insulin clearance assuming no dialysis, and a lower insulin resistance. |
| Step 2:<br>Data-based Profiles with Laboratory Measurements | Using the first three days of laboratory measurements of patients in phenotype G8, we observe significantly higher AST values compared to G9 ($p<0.001$), G11 ($p<0.001$), and G12 ($p<0.001$). This indicates higher level of dysfunction in liver and kidney in phenotype G8 compared to all other phenotypes except G10. We also observe that patients in phenotype G8 are missing laboratory measurements of AMY.<br>Using the last three days of laboratory measurements of patients in phenotype G8, we observe significantly higher AST values compared to G9 ($p<0.001$), G11 ($p=0.004868$), and G12 ($p<0.001$). This indicates different liver functionality in phenotype G8 compared to all other phenotypes. We also observe that patients in phenotype G8 are missing laboratory measurements of LIPA and BUN. |
| Step 3:<br>Data-based Profiles with ICD 9/10 Data | Based on ICD 9/10, patients associated with ICU stays in phenotype G8 were diagnosed as having percentage counts of hyperglycemia or sepsis 67% of the time during the complete stay, and 11% in the last three days. Patients associated with ICU stays in phenotype G8 were also diagnosed as having percentage counts of dialysis or renal/kidney failure 78% of the time during the complete stay, and 22% in the last three days. |
| Step 4:<br>Concordance Between Model-based Phenotype and Data-based Profiles | Only through ICD 9/10 data can we know that phenotype G8 had severe kidney dysfunction and a high incidence rate of hyperglycemic events or sepsis at the ICU admission. Both model and laboratory measurements indicate liver dysfunction in phenotype G8 in the last three days. Both laboratory measurements and ICD 9/10 data indicate patients in phenotype G8 managed to alleviate kidney dysfunction in the last three days. Only through ICD 9/10 data can we know that patients in phenotype G8 managed to alleviate hyperglycemia and even sepsis approaching ICU discharge. |
| Step 5:<br>Synthesized Phenotypic Description | Patients in phenotype G8 had a slightly decreased pancreatic insulin secretion, much higher insulin clearance assuming no dialysis, and a slightly decreased insulin resistance in the last three days. They had liver dysfunction, severe kidney dysfunction, and a high incidence rate of hyperglycemic events or sepsis at the ICU admission, but managed to alleviate kidney dysfunction and hyperglycemia, even sepsis approaching the ICU discharge. |

#### 3.3.5 Quantitative Clinical Face Validity of Clinical Mapping Accuracy and Consistency

We used face validity [58] to quantitatively evaluate the pipeline’s practical clinical concordance based on an endocrinologist’s review of patient notes in the last three-day period. We opted to validate qualitative discrete phenotypes for each patient at the level of discretized physiological descriptors: the associated physiological interpretations (cf. Sec. 3.2.4) of parameter triplet (*a*_1_, *t_p_*, *R_g_*) estimation. This patient- and parameter-specific quantitative approach was similar to but enriched from the qualitative customized ranking method that produced discrete phenotypes. In other words, here we prescribed percentage likelihoods of parameter estimates binning into levels (low, medium, and high) from associated EDFs as weights, where the levels were relative to the cohort range. Combining the weight-based binning with physiological interpretation, for each patient and each physiological descriptor estimate of the patient, we statistically analyzed the similarity between expert reading of patient health and interpretable binning expectation based on one minus a ordinal distance metric.

For 20 ICU stays picked from the resolved set of 45 ICU stays in cohort B3, the interpretable binning expectations quantitatively agreed with associated physiological functionalities that were identified by the blind chart review for 20 patients associated with these ICU stays. The concordance–measured by the mean of 20 responses–of each parameter were: 83% *±* 27% for *a*_1_ (insulin secretion), 66% *±* 35% for *R_g_* (insulin resistance), and 52% *±* 44% for *t_p_* (insulin clearance), respectively. However this concordance might be confounded by interventions and therapies in ICU that are not estimable from the model. For example, the dialysis treatment due to acute kidney disease confounded our estimation of insulin clearance, leading to a lower concordance of model parameter *t_p_*. Table 5 summarizes the potential confounding factors that might affect model estimation of insulin clearance, combining nurses notes during patient ICU stay and clinician notes during face validation, which was further discussed in conclusion Sec. 4.4.

**Table 5:** Confounding factors of insulin clearance from clinician perspective and ICU notes that might affect face validity concordance. 15 of 20 picked ICU stays either had hemodialysis treatment or continuous veno-venous hemofiltration (CVVH), or had kidney failure related conditions indicating the necessity of dialysis treatment during the ICU stay. Diagnoses based on ICD 9/10 codes had dagger superscripts †. The asterisk superscripts * indicated diagnoses, therapies, and procedures identified from free text narratives.

| Clinical Confounding Factors in Face Validation (FV) |  |  |  |  |
| --- | --- | --- | --- | --- |
| Patient Number | Clinician Perspective | Diagnosis | Therapy/Procedure (FV period) | Therapy/Procedure (complete ICU stay) |
| 1 | passed away on the last day after withdrawal of support |  |  |  |
| 2 | passed away on the last day after withdrawal of support | other acute kidney failure<sup>†</sup> |  |  |
| 3 | passed away on the last day after withdrawal of support |  |  | placement and removal of hemodialysis access non-tunneled double lumen dialysis catheter |
| 4 |  | CKD (chronic kidney disease) stage 2<sup>†</sup>, renal insufficiency* |  | placement and removal of hemodialysis access non-tunneled double lumen dialysis catheter, heart transplant* |
| 5 | CVVH | renal failure acute (HC code)<sup>†</sup> |  |  |
| 6 | previous lung transplants, CVVH |  |  |  |
| 7 | T1DM | acute kidney failure unspecified<sup>†</sup> |  |  |
| 8 |  |  | placement and removal of hemodialysis access non-tunneled double lumen dialysis catheter |  |
| 9 | from low blood pressure to then improved renal function |  |  |  |
| 10 | heart transplant |  |  |  |
| 11 | RRT hemodialysis |  |  |  |
| 12 | previous heart transplants |  |  |  |
| 13 | worsening renal function |  |  |  |
| 14 |  | medical renal disease* |  | renal ultrasound* |
| 15 |  | chronic renal failure* |  |  |

### 3.4 Additional Insights: Impact of Health Care Process on Phenotype Labeling and Clinical Concordance of Discrete Phenotypes

To account for the unresolved biases in our phenotype computation arising from health care process, e.g., hemodialysis or insulin interventions, we altered confounding factors that perturb physiological functionalities. Specifically, because the model only includes physiological mechanics, its inability to differentiate unmodeled intervention effects or incorrect model assumptions from modeled physiological mechanics can reduce phenotype accuracy. We performed this natural experiment of perturbing the pipeline by removing T1DM patients with abnormal insulin production deficiency that is not well represented by model assumptions of typical endogenous blood-glucose management. We removed patients with T1DM–eight in cohort B, seven in cohort B3, and two in the face validation cohort–and re-ran from scratch the phenotype computation, providing additional insights into our hypothesis.

Table 6 shows machine sensitivity to phenotypic labeling in terms of predictive power, revealing the impact of health care process on phenotype computation. Table 6 top two colored sub-tables detail updated phenotypic memberships from phenotypes G8-G12 (cohort B3) to four coherent phenotypes (cohort B3 excluding T1DM) using the same pipeline configuration. Table 6 bottom table details the comparison of phenotype-specific predictive power. Removing some of the unresolved bias improved the pipeline’s internal evaluation. Additionally, phenotype sensibility and clinical relevance at the physiological descriptor level were also improved during face validation for all three physiological descriptors binning: from 83%*±*27% to 87%*±*26% for insulin secretion, from 66% *±*35% to 71%*±*33% for insulin resistance, and from 52%*±*44% to 58%*±*43% for insulin clearance, respectively.

**Table 6:**
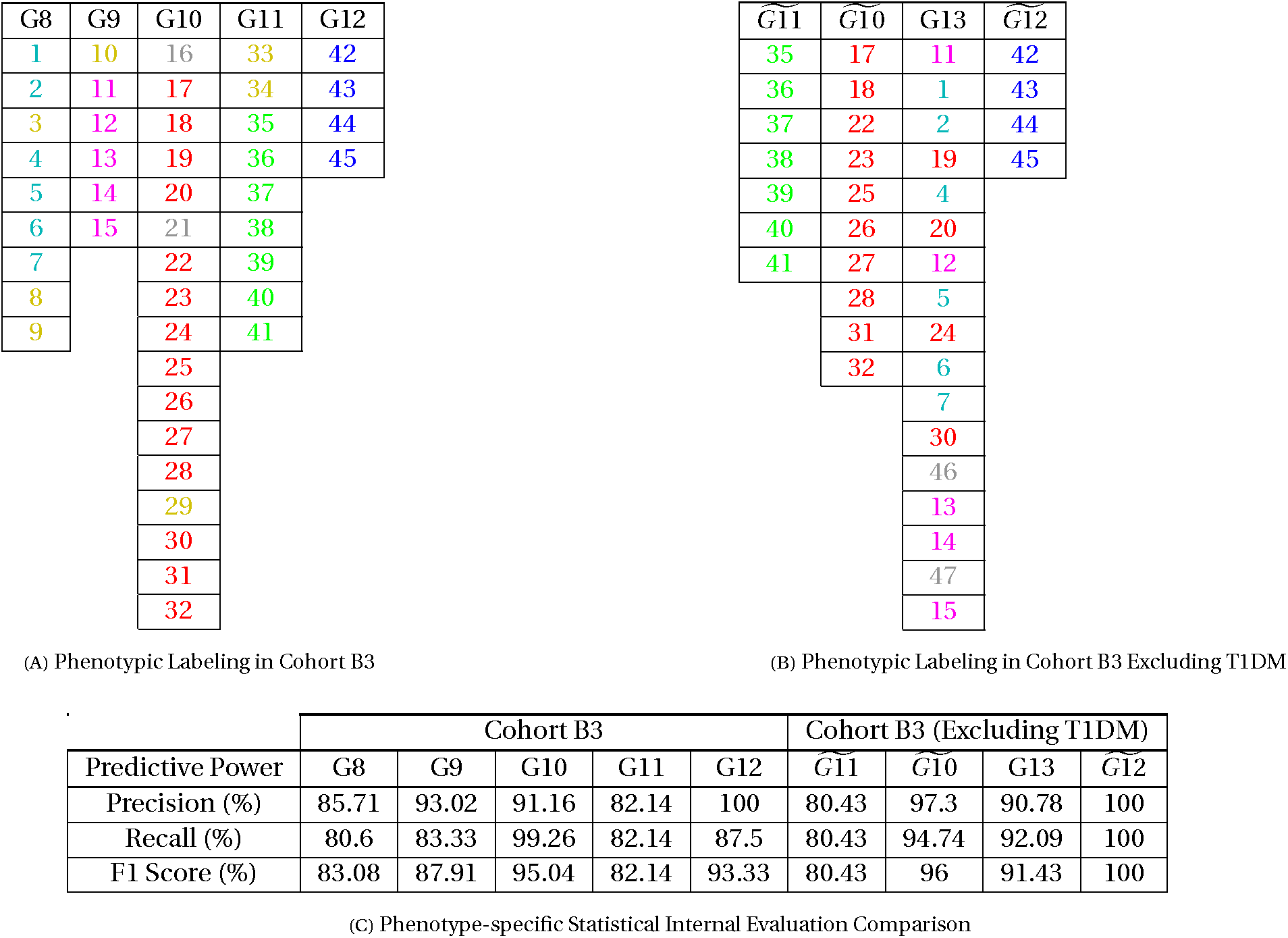
The impact of health care process on machine sensitivity to phenotypic labeling. Two sub-tables show colored tracking of patient phenotypic memberships from **(A)** phenotypes including T1DM patients to **(B)** updated phenotypic labeling excluding T1DM patients in cohort B3, columned by phenotypes in each cohort, where patients were indexed without ranking. The color scheme was consistent with Figures 4 and 5 to indicate each patient’s association with phenotypes G8-G12, which included seven T1DM patients highlighted in yellow. Patients in grey were analyzed in the current cohort but dropped from the other cohort as we considered them to be associated with SVC-identified auxiliary phenotypes below a sufficient sample size (cf. Sec. 2.3.4). This updated labeling reorganized phenotype identification by still keeping the original phenotypes G11, G12, and the majority of patients in G10. The new phenotype, ‘G13’, included all patients previously labeled as phenotypes G8 and G9 and a portion labeled as G10. This phenotype shared characteristics of G8 and G9 in terms of alleviated kidney dysfunction and similar levels of insulin secretion and resistance. The splitting of G10 among two phenotypes related to liver problems and kidney disease reflected by higher levels of aspartate aminotransferase, blood urea nitrogen, and creatinine in patients later clustered into ‘G13’. We included in the Appendix (Sec. A.6) the updated parameter estimates grouping and phenotypic labeling process via t-SNE and SVC. Sub-table **(C)** is the comparison of phenotype-specific predictive power across all three performance metrics: precision, recall, and F1 score. The average predictive power was also improved in accuracy (from 89.39% to 91.97%), macro F1 score (from 88.30% to 91.59%), and weighted F1 score (from 89.26% to 91.58%).

## 4 Conclusion

### 4.1 Key Results and Related Contribution to Current Phenotyping Methods

#### 4.1.1 Summary: The Three-stage Methodological Pipeline

In this paper we developed a novel DA-driven methodological pipeline that compute personalized, physiologically-anchored, time-specific, and interpretable phenotypes from EHR data. We applied the pipeline to ICU patients using an established mechanistic model–the ultradian model–and MCMC inference to extract unmeasurable physiological properties using blood glucose measurements and administered nutrition and insulin. Much of the phenotype computation was performed on a cohort of 109 ICU stays from 109 patients with a focus on the last three-day period of enteral nutrition in the ICU and with the further restriction to patients who were given either no or short-acting insulin. Working within the inverse problem framework, we selected ultradian parameters for inversion and estimation based on objective criteria to avoid technical complications arising from non-unique inference solutions. Estimating these parameter balanced model flexibility and fidelity, which was challenged by physiological non-stationarity and data sparsity for individual-level modeling. This estimation process led to individual ICU stay to be characterized by qualified continuous estimates of selected model parameters in probability distributions. Through this representation process we inferred and created new physiological information that we then clustered into cohort-scale labeled phenotypes characterized with continuous distributions, which were subsequently discretized and validated.

The phenotype computation was performed using unsupervised ML methods including t-SNE for dimensionality reduction, and SVC for clustering parametric representations of ICU stays in the cohort into groups of labeled continuous-space phenotypes that were physiologically-anchored. Then a four-step process extracted the physiological meanings of these labeled phenotypes by reconstructing physiological information from SVC-identified clusters based on physiological descriptors of the estimated parameter triplet (*a*_1_, *t_p_*, *R_g_*). This process also rendered the discrete forms of continuous physiological phenotypes, defined as discrete phenotypes, as proxies of model-inferred physiological functions for statistical analysis, qualitative clinical interpretation, and quantitative external validation in a lower dimensional space. We found six coherent discrete phenotypes in the first three-day, six in the middle three-day, and five in the last three-day period. Phenotype interpretation leveraged the physiologically-defined model parameters that encoded clinically important but unmeasurable endocrine functions. Based on these new interpretations, we also generated a hierarchy visualization to assist contextual interpreting and delineating discrete phenotypes for clinical mapping.

The phenotype evaluation and validation was imperative in examining the pipeline machinery and establishing the potential clinical utility. Computed discrete phenotypes were quantitatively evaluated for predictive power and externally validated for clinical coherence and consistency. Specifically, clinically observed inter-phenotype difference in external data including selected laboratory measurements and ICD 9/10 codes were quantitatively consistent with the five discrete phenotypes identified in the cohort of the last three-day period. Quantitatively, we further used face validity to check the pipeline’s clinical concordance at the level of three model physiological descriptors. Arguably the most clinically impactful parameter to establish clinical translatability was insulin secretion (*a*_1_) that had the highest clinical concordance of 83%*±*27% based on face validity, while weaker results were found for insulin resistance (*R_g_*) and insulin clearance (*t_p_*) that were influenced by potential confounding factors.

We can construct time-varying phenotypes computed over a sequence of data assimilation windows to produce time-specific physiological phenotypes that are estimated by assuming stationary physiological functions within a specific time window. There also are existing efforts in the temporal phenotyping domain that identify patterns or classify evolution in health care process from longitudinal EHR through sequential pattern mining [59][60][61], phenotypic topics identification [62], or graphical-based models [63]. Such methods leverage conventional ML, deep learning, and statistical analysis within patients’ population strata. We did not include an empirical comparison of our method with these other temporal phenotyping methods for two primary reasons: (i) Our narrowly-specified physiological phenotypes are defined with individually-estimated and data-inferred unmeasurable physiological properties (viz. insulin secretion, clearance, and resistance) that cannot be compared directly with existing EHR information. Our evaluations, which compare ICD 9/10 data, lab traits, and clinical notes to model-derived physiological properties, are therefore designed to assess comprehensively whether these contextually narrow and unmeasurable phenotypes are cohesive and clinically coherent. (ii) The temporal phenotyping methods are generally ML-dependent and require existing or extractable phenotypic labels for training and testing. As mentioned in Sec. 2.4, such physiologically specific labels for insulin secretion, insulin clearance, and insulin resistance are not present or easily computable from EHR data. As a result, most temporal phenotyping methods are unable to estimate or implicitly identify the same unmeasurable and continuous endocrine properties. Understanding these differences is important. While many current temporal phenotyping methods detail how health care process variables such as laboratory measurements vary, the methodological pipeline we present here is narrowly focused on phenotypic information representing particular physiological mechanisms. Because of these reasons, the features that make up these two methodological phenotype definitions are incompatible for a direct comparison. Further, the evaluations in our narrowly-specified physiological context and the evaluations in the other temporal phenotyping contexts are too different to render meaningful comparisons. Nevertheless, there is compelling potential to integrate our methods into other methods. Our methodology could supply additional physiological information that is not explicitly present in the EHR into existing temporal representation mining methods. Incorporation of clinically interpretable, patient-specific, continuous, and time-specific parameter estimates of, e.g., insulin secretion, may expose new features within sequential health care patterns as well as improve disease prediction.

Finally, we excluded phenotype utility evaluation from this paper, which requires the construction of an expert panel to evaluate how computer-generated phenotypic information would affect clinician understanding or decision-making. Establishing usefulness is a complex qualitative analysis problem that demands massive data collection and analysis for clinician interviews. While this problem is an interesting area of our future research, we left it out of the scope of this paper evaluating the computation of unmeasurable physiological properties from EHR.

#### 4.1.2 Pathway for Automating and Embedding the Pipeline into a Health System

The mechanical computational parts of this largely automated pipeline system can easily scale to high-throughput in real-time settings, as we can increase the cohort size or simply use more CPUs for higher fidelity cluster results based on the automation. Necessary manual tuning and optimization procedures kept in the pipeline inject expert knowledge in helping with cohort exclusion criterion, deciding the model complexity, making adjustments to default method configurations, and identifying a plausible number of phenotypes. In tandem with the systematic automation, we also inadvertently touched on the pipeline’s generalizability as we computed phenotypes with shared methodological choices for two ICU patient data sets across two hospitals.

### 4.2 Methodological Choices

We made several preliminary methodological choices, including picking a DA model, parameter estimation techniques, dimension reduction approaches, and clustering methods, and our methodology can presumably be sensitive to these methodological choices.

#### 4.2.1 Physiological Model Choice: Fidelity and Flexibility

Which model we pick and the model complexity are impactful to what phenotypes are possible, computable, and with different resolutions. The mechanistic physiological model we pick is only an approximation and hypothesis-based representation of the actual system, as a model reduces nature to several elements found to be adequate to explain a particular phenomenon. On one hand, a very simple model may fail to represent the EHR data hence harm the glycemic states prediction precision. At the other extreme, a model with higher complexity will require high resolution data to estimate that normally do not exist in clinical settings, leading to model identifiability issues with more vexing model accuracy. This means that even if we have the correct data available, different models may provide different explanations for the same physiological process, e.g., the current model applied to T1DM patients, which can be differently inaccurate or wrong.

To this end, accurate and robust phenotyping requires care when selecting parameters of a model that represents the system with a balanced physiological fidelity and flexibility. We picked the mechanistic ultradian model and only selected subsystem parameters to estimate for phenotype computation in this paper, as we were posed with more severe data sparsity due to accommodating the ultradian model for an ICU setting with simplified pharmacodynamics only with short-acting insulin. Additionally, because the cohort we computed phenotypes from has mixed patients with T1DM and non-T1DM, excluding T1DM individuals whose physiological characteristics do not adhere to model assumptions may be beneficial to phenotyping accuracy. Nevertheless, as other data sources become available, e.g., continuous glucose monitors, other more complex models may be more advantageous. We were also not exhaustive in our use of different models, albeit we suspected that in the ICU context one important requirement is the model’s ability to exhibit glycemic oscillations.

#### 4.2.2 Alternative Methodological Choice Example in Dimension Reduction of Parameter Estimates: t-SNE vs PCA

An alternative approach of dimension reduction in the phenotype computation stage is the PCA method that can preserve variation with distribution compression. However, its usage is inadequate for the purpose of phenotyping as multicollinearity due to model parameter synergy necessitates combining all EDFs associated with the same patient, which complicates cohesive phenotype computation that we expect to be independent of patient label.

### 4.3 Motivation for Identifying Non-mutually Exclusive Phenotypes

We found that the SVC-identified coherent phenotypes (cf. Sec. 3.2.4) were not always mutually exclusive, meaning that ICU stays clustered into different phenotypes shared similarities in terms of model-estimated physiological properties. There are two possible reasons for this to happen, even when our methodology indeed computes the right phenotypes. One main reason is that our method allows for overlapping phenotypic characteristics by representing an ICU stay as a continuum in methodological parts. Secondly, the non-mutually exclusive phenotypes reflect the balance between biological fidelity and model flexibility, where we maximize the likelihood of having a unique model solution using a flexible model. When it is the case that we use incorrect methodological choices in computing phenotypes, the chosen mechanistic model may be simpler than required or we have inaccurate estimates due to inappropriate MCMC method choice.

### 4.4 A Deeper Dive into Face Validity

Measuring the concordance between model-based phenotypes and clinical diagnosis at the phenotype level through face validation can be problematic. The clinician might have a relatively low level of confidence in categorizing the patients, as a patient might only satisfy certain aspects of the descriptions. Therefore, we disentangled this complex problem by validating estimates of physiological descriptors rather than phenotypes.

We had varying concordance among parameter estimates. Referring to Table 1 and section 3.3.5 with Table 5, the decreasing concordance with increasing uncertainty for binning physiological descriptors in sequence from *a*_1_ (83% *±* 27%) to *t_p_* (52% *±* 44%) was probably due to an increasing amount of health care interventions and other confounding factors. Nevertheless, we hypothesize that the accuracy of recorded insulin dosage time series in EHR data enabled us to identify model parameter *R_g_* that relates to insulin resistance.

The validation process at the parameter level, though, is still complicated by translation issues of across-domain gap and language disconnect. The discordance in delineation and quantification between knowledge domains, e.g., our definition of the average glycemic states in ICU is different from a typical clinician mindset, implies an unaddressable across-domain gap that is difficult to detect and also a less related topic in this paper. The disconnect between the language of physiology and language of clinical diagnosis, e.g., the mapping between physiology and SNOMED CT can lead to error and bias. This complex translation problem might be treatable through the framework of human-computer-interaction (HCI) for future clinical decision support, where related work can motivate research in cognitive science, behavioral science in biomedical informatics, etc., that mainly hinge on users’ intention of usage [64].

## Data Availability

All data produced in the present work are PHI ruled by HIPPA and unavailable for sharing.

## Acknowledgments

The work was supported by grants from the National Institutes of Health R01 LM006910 “Discovering and applying knowledge in clinical databases,” and LM012734 “Mechanistic machine learning,”.

## A Appendix

### A.1 The Ultradian Model Mathematical Formulas

The mechanistic ultradian model has six ordinary differential equations with three physiological states (*I_p_* is plasma insulin, *I_i_* is remote insulin, *G* is plasma glucose) and a three stage filter (*h*_1_, *h*_2_, *h*_3_) that represents how plasma insulin affects glucose.

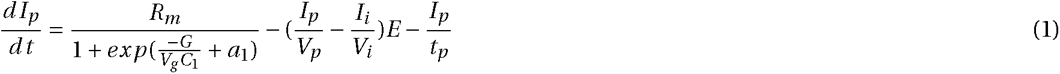

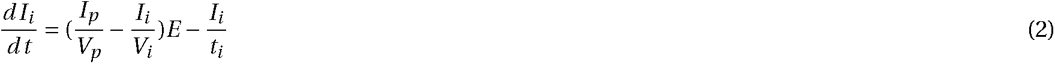

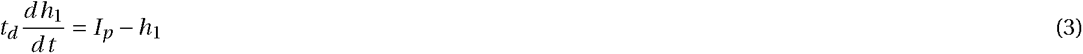

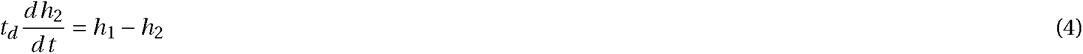

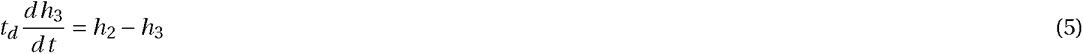

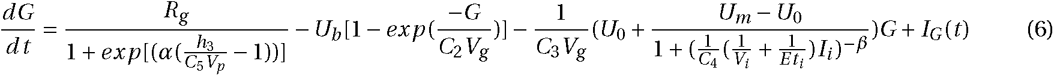

### A.2 The Houlihan Algorithm for Model Parameter Selection

The Houlihan algorithm uses a specific combination of feature metrics and influence function–lasso in L1 regularization, L2 regularization, and principal component analysis (PCA)–as a parameter selection method to rank order single parameter and parameter pairs/triplet [41]. Under the Houlihan approach, parameters *a*_1_, *t_p_*, and *R_g_* in ultradian model [22] were found to have the highest-rank individual influences on model fidelity, while parameter pairs of (*a*_1_,*C*_1_), (*R_g_*,*C*_3_) and triplet of (*a*_1_,*C*_1_, *t_p_*), (*a*_1_,*C*_1_,*C*_3_) were also ranked high. The method also justified reducing model parametrization to only a few core parameters set nominees: (*a*_1_, *t_p_*, *R_g_*, *C*_1_), (*a*_1_, *t_p_*, *R_g_*,*C*_3_) and (*a*_1_, *t_p_*, *R_g_*) became our model parameter selection nominees, noting that both (*a*_1_, *t_p_*, *R_g_*) and (*C*_1_, *t_p_*, *R_g_*) correspond to representing the kidney-liver-pancreas subsystem. Other parameters besides *a*_1_, *t_p_*, *R_g_*, *C*_1_, and *C*_3_ are likewise physiologically interpretable, but in this paper we focused only on these parameters with the highest influence.

### A.3 Geweke Statistic Fractions Identification

We used Geweke statistic as one of the objective quality criteria to screen Markov chains for intra-chain convergence under limited computing resources. The convergence in Geweke statistic is assessed by comparing the initial and terminal fractions of a Markov chain. Since we normally start our iterations somewhere not close to the true value, for a converged chain we expect to see distinctive difference between the terminal and initial portion of iterations, which indicates some degree of convergence approaching the tail of iterations. We only kept chains with significant p-values in Geweke statistic under the significance level *α*=0.06. Although Geweke suggested using 10% and 50% as the initial and terminal fractions, it is not a burden grid searching for the best in a careful tuning exploration. We explored in Figure A.1 different values for the initial fraction while fixing the terminal fraction to be a relatively large ratio, as we would like to only keep chains that converge and also stay stable at the tail. We identified the best Geweke fractions as 3% for the initial and 50% for the terminal iterates.

**Figure A.1.**
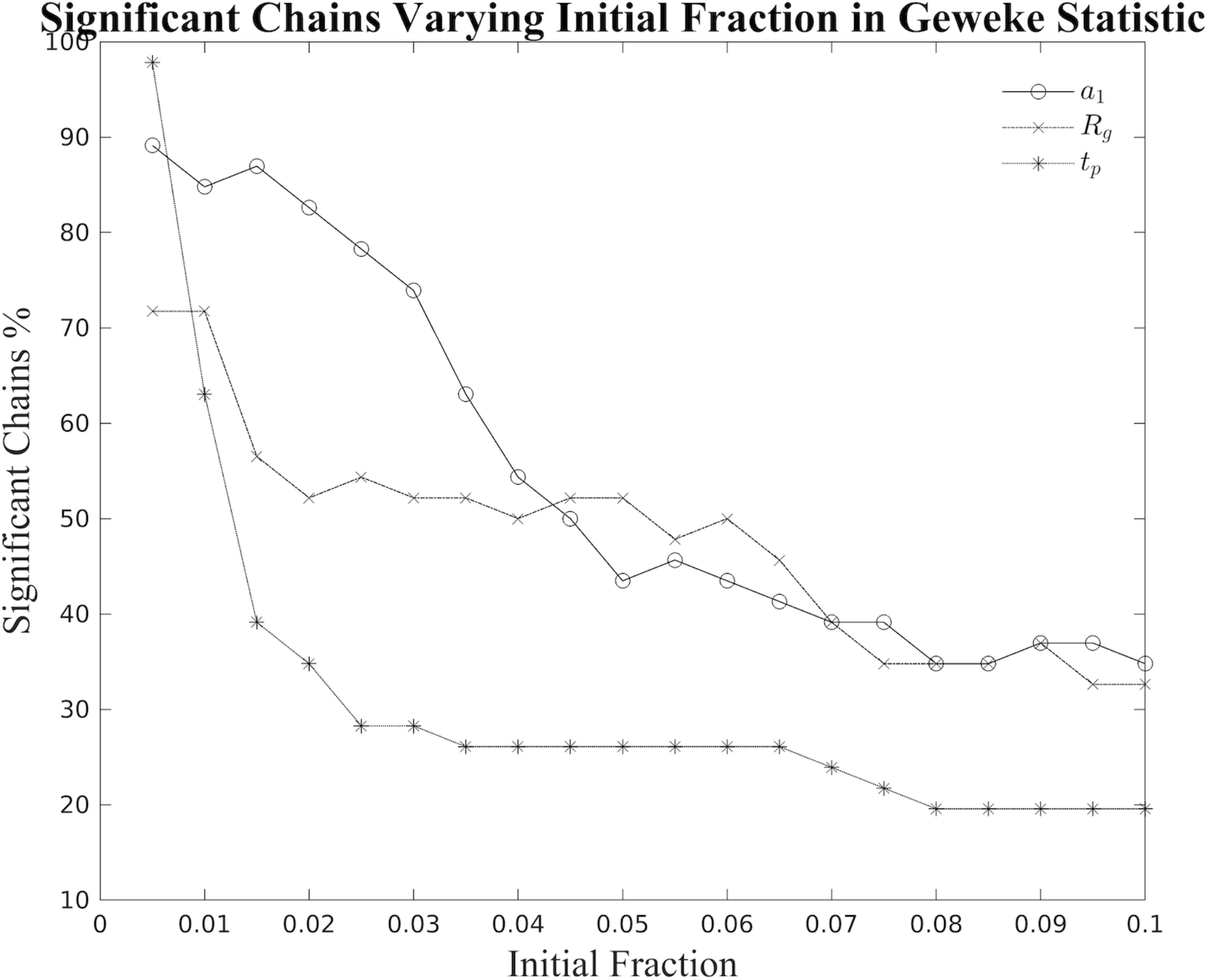
: Fraction identification of Geweke Statistic. Using cohort A data, we ran 3 Markov chains for the parameter triplet (*a*_1_, *R_g_*, *T_p_*) then counted percentage of converged chains for each parameter using Geweke statistic with the initial fraction ranging from 0.05 to 0.1 and the terminal fraction fixed at 0.5. We chose the initial fraction as the first smallest value that decreased the convergence percentage for all parameters, hence we identified the initial fraction as 0.03. With this initial fraction, 47 out of 90 chains converged in Geweke statistic. Chains selection optima from cohort A were adopted for cohort B.

### A.4 Customized Ranking of Model Parameters for Interpretation

Based on the estimation of the parameter triplet (*a*_1_, *t_p_*, *R_g_*), the customized ranking method compares each group’s posterior mean to the posterior mean across all groups identified in a specific window, e.g., the last three days of the complete ICU stay. Then setting the posterior mean across all identified groups as the midpoint, group mean that falls in (0,1/3*midpoint) was ranked as ‘much lower’ and noted as ‘*↓↓↓*’; (1/3*midpoint, 2/3*midpoint) was ‘moderately lower’ and ‘*↓↓*’; (2/3*midpoint, midpoint) was ‘slightly lower’ and ‘*↓*’; (midpoint, 4/3*midpoint) was ‘slightly higher’ and ‘*↑*’; (4/3*midpoint, 5/3*midpoint) was ‘moderately higher’ and ‘*↑↑*’; (5/3*midpoint, 2*midpoint) was ‘much higher’ and ‘*↑↑↑*’.

### A.5 List of Laboratory Measurements and ICD 9/10 Codes for External Validation

The list of ICD codes we picked includes keywords of ‘hyperglycemia’, ‘sepsis or septicemia’, ‘renal failure or kidney failure’, ‘renal’, and ‘dialysis’, which indicate problems with organs whose functions are encoded in model parameters. We tabulated in Table A.1 eight selected laboratory measurements that are related to pancreas, kidney, and liver functions in general as clinically observable evidence for external validation. These laboratory measurements are clinical dysfunction indicators of certain organs relatable to the extracted physiological properties. Noting here that we only selected laboratory measurements not used in phenotype computation for external validation. For example, we did not select glucose laboratory measurements in terms of hyperglycemia for external validation, because we used glucose in the phenotype computation as required by the model estimation.

**Table A.1:** Selected laboratory measurements as external data and their related physiological functions encoded in model parameter triplet (*a*_1_, *t_p_*, *R_g_*).

| Selected Laboratory Measurements in EHR for External Validation |  |
| --- | --- |
| Laboratory Measurements | Related Physiological Function |
| Amylase (AMY) | Pancreatic enzyme production for digestion (an indicator of insulin secretion) |
| Aspartate aminotransferase (AST) | Liver problems |
| Lipase (LIPA) | Pancreatitis and other pancreatic disorders |
| Creatinine (CREAT) | Kidney filtering capability in general (an indicator of insulin clearance under a non-dialysis condition) |
| Blood urea nitrogen (BUN) | Kidney filtering capability in general (an indicator of insulin clearance under a non-dialysis condition) |
| Alanine aminotransferase (ALT) | Signs of jaundice indicating liver/kidney dysfunction |
| Albumin (ALB) | Part of a comprehensive metabolic panel of liver and kidney |
| Bilirubin (BIL) | Liver damage or disease |

### A.6 Updated Phenotype Projection Excluding T1DM Patients

Referring to Section 3.2.3, this section presents in Figure A.2 the updated visualization of parameter estimates projection in the 2D t-SNE space and the further clustering into mechanistically distinctive phenotypes with SVC, for cohort B3 without T1DM patients.

**Figure A.2:**
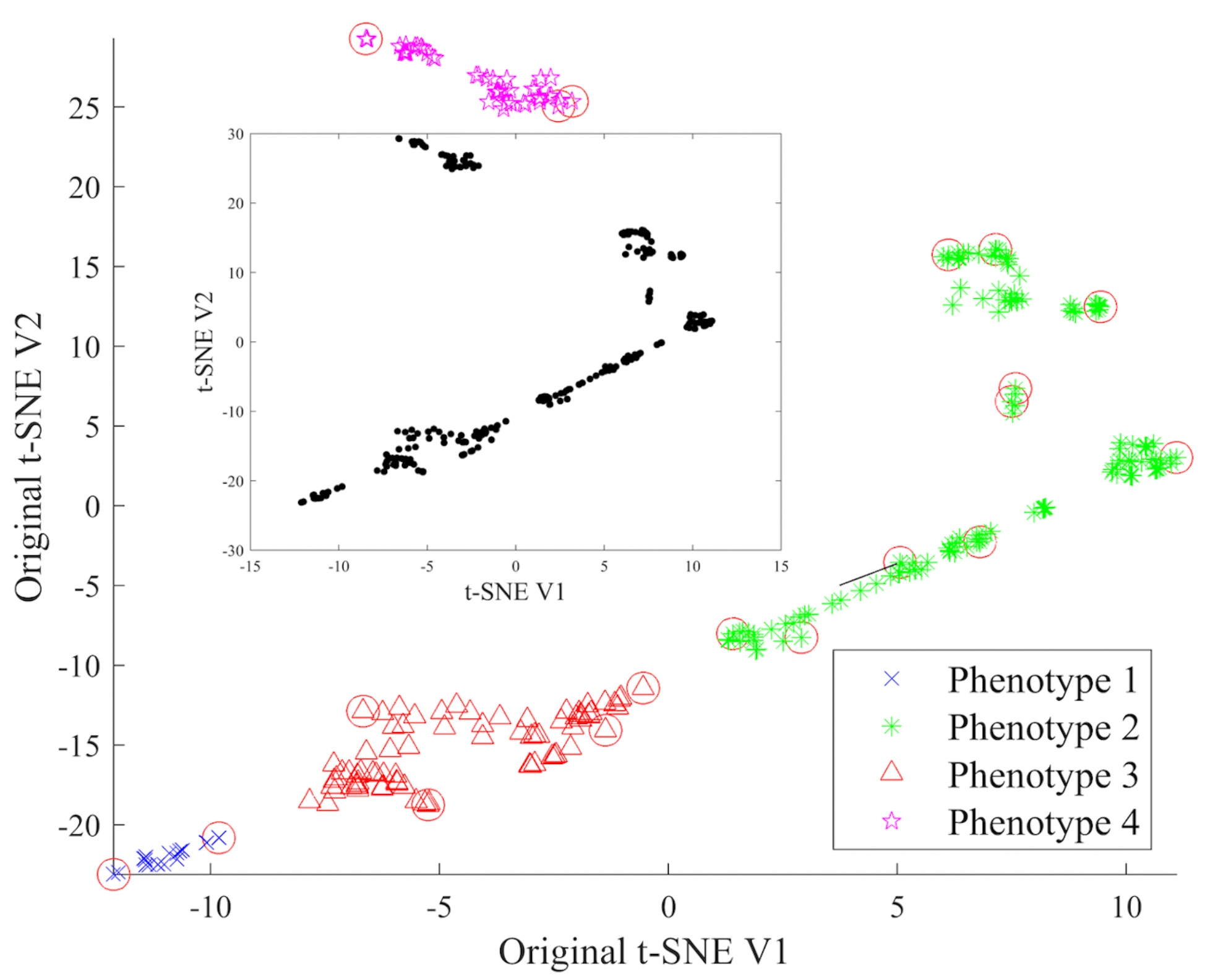
**Step 1 (inset)**: Raw and un-clustered t-SNE dimension reduction of selected chains summary of 38 ICU stays in cohort B3 excluding T1DM using the same methodological choices. **Step 2**: Phenotype computation with SVC that clusters t-SNE coordinates of chains summaries. Circled coordinates are support vectors that lie on the feature space sphere boundaries. Four clusters were identified indicating four potential phenotypes.

